# Can we detect the undetected? Comparing the prodromes of individuals with first episode psychosis detected and undetected by clinical high risk for psychosis services: an electronic health record study

**DOI:** 10.1101/2025.06.26.25330305

**Authors:** Maisie Inns, Madison Gill, Maite Arribas, Yanakan Logeswaran, Amedeo Minichino, Thomas J. Reilly, Andrea De Micheli, Jai L. Shah, Philip McGuire, Paolo Fusar-Poli, Dominic Oliver

## Abstract

**Background and Hypothesis:** The majority of first episode psychosis (FEP) patients are undetected (DET-) by clinical high risk for psychosis (CHR-P) services prior to onset and therefore do not receive preventive care for psychosis. We compared features of the psychosis prodrome (symptoms and substance use) between DET- and FEP patients detected by CHR-P services (DET+) to determine whether they share a common prodromal phase.

**Study Design and Measures:** Retrospective RECORD-compliant electronic health record cohort study. We extracted 65 prodromal features (symptoms and substance use before FEP onset) using natural language processing to assess the presence, duration and first presentation of the psychosis prodrome and occurrences of features across the prodrome. Duration and feature occurrences were compared between DET+ and DET-individuals using Mann-Whitney U tests and Wilcoxon Effect Size, while presence and first presentation were compared using logistic regression.

**Study Results:** 1,545 FEP patients (n=119 DET+ [mean age 28.7 years; SD=9.4; 61.6% male]) were included. There were no significant differences in the presence (DET+=85.0%, DET- =85.6%, p=0.83) or duration (DET+=18.8 months, DET-=18.4 months, p=0.89) of the psychosis prodrome. There were no significant differences in first presentation of psychotic symptoms between groups (p_corr_>0.05). Frequency of occurrences of thought broadcasting (r=0.07, p_corr_=0.04) was higher and hostility (r=0.08, p_corr_=0.04) lower in DET+ compared to DET-across the prodrome, though effect sizes were small.

**Conclusions:** DET+ and DET-individuals experience similar psychosis prodromes prior to FEP onset. DET-individuals can likely be identified earlier if detection strategies are improved.

## Introduction

Early detection through the clinical high risk for psychosis (CHR-P) state is the most promising strategy to delay or prevent the onset of a first episode of psychosis (FEP)^1,2^. Currently only a minority of FEP patients (5-13%)^3–5^ are being detected by CHR-P services before the onset of the disorder, highlighting a need to improve current detection strategies.

78.3% of FEP patients experience a prodromal phase^8^. This is variably-defined, including psychotic symptoms but also nonspecific psychotic symptoms that are contiguous into the emergence of FEP^8^, and hetereogeneous with estimates across individual studies ranging from 22-100%^8–11^. Current initiatives to improve detection of individuals at risk depend on the assumption that those currently undetected prior to FEP onset experience a detectable prodromal phase^6^ that is similar to those experienced by people currently detected by CHR-P services, typically characterised by attenuated positive psychotic symptoms. These prodromal symptoms are assessed by validated psychometric instruments, such as the Comprehensive Assessment for At Risk Mental States (CAARMS) or the Structured Interview for Psychosis risk Syndromes (SIPS), which have excellent population-level prognostic accuracy^7^. If those undetected prior to FEP do experience a prodromal phase that is similar to a CHR-P phase, we would need to improve detection strategies so that CHR-P assessments are conducted earlier and reach a wider population. Alternately, if these prodromes are different from the CHR-P phase, then alternative detection strategies need to be developed outside of the CHR-P paradigm. Understanding how prodromal symptoms first present and develop in people detected and undetected by CHR-P services is thus instrumental for guiding future preventive strategies^10,12,13^.

Our understanding of the prodrome to FEP is predominantly based on studies assessing prodromal symptoms retrospectively by interviewing individuals with FEP. While these studies are informative, they may be influenced by recall bias, only provide information about a select group of symptoms at a single time point, and may not be directly generalisable to real-world clinical practice. Similarly, while prospective CHR-P studies address some of these issues, as those who transition represent a minority of the FEP population^3–5^, they are not truly representative. Electronic health record (EHR) data can potentially address these limitations as it is collected more contemporaneously from a representative clinical sample (i.e. all FEP patients) and can be combined with natural language processing (NLP) algorithms to capture real-time (including prodromal) symptoms across a longer time course from free text in clinical notes and letters.^14^

Using EHR data, we aimed to primarily compare the (i) presence, (ii) duration and (iii) first presentation of the psychosis prodrome and (iv) mean number of occurrences of psychotic symptoms across the prodromal period in individuals detected (DET+) and undetected (DET-) by a CHR-P service prior to FEP onset. As a second step, we compared the (i) duration, (ii) first presentation and (iii) mean number of occurrences of all symptoms and substance use features (overall prodrome) between the two groups.

## Methods

This study adhered to the Reporting of Studies Conducted Using Observational Routinely Collected Health Data statement (RECORD) (eTable 1).

### Setting

Data was sourced from South London and Maudsley National Health Service (NHS) Foundation Trust (SLaM). SLaM consists of four boroughs in South London (Lambeth, Southwark, Lewisham, and Croydon) and provides the largest range of NHS Mental Health Services in the UK, covering 1.3 million people^15^. The incidence of psychosis within SLaM is one of the highest worldwide, with reports estimating between 58.3-71.9 cases per 100,000 compared to the national average of 41.5 cases^16^.

This study focused on a comparison between patients with FEP who had been detected (accepted referral) by OASIS (Outreach and Support In South London) prior to disorder onset (DET+) and those who had not (DET-). OASIS is a specialist multidisciplinary community mental health service within SLaM for individuals at CHR-P^17^. OASIS includes individuals within a catchment area of 443,050 people aged between 14-35 years (matching the age range of most CHR-P individuals), and has a yearly caseload of approximately 140 CHR-P individuals^16,17^.

EHRs have been utilised across all SLaM services since 2006. Full, anonymised clinical data from the EHRs has been made available for searching, retrieval and analysis through the Clinical Record Interactive Search (CRIS). CRIS is approved by the Oxfordshire Research Ethics Committee C (Ref: 23/SC/0257) and is extensively validated^18–20^. Participants were able to opt out of having their data utilised for research through SLaM’s opt out scheme^21^.

### Study design and population

This research utilised a retrospective (up to 12 years), real-world, EHR-based, RECORD-compliant, cohort study design (eTable 1). All individuals who were accepted as referrals to early intervention for psychosis teams in SLaM between 1^st^ January 2008 and 10^th^ August 2021 who received a primary ICD-10 diagnosis of a non-organic psychotic disorder (eTable 2) were eligible for inclusion. The date of FEP onset (index date; T-0 months) was defined as the earliest date from: i) first ICD-10 diagnosis of a psychotic disorder recorded in the EHR; ii) accepted referral to the early intervention team; iii) date of hospital admission where an ICD-10 diagnosis of a non-organic psychotic disorder was recorded at discharge; or iv) prescription of an antipsychotic medication equal to or greater than the minimum FEP dose defined by the 14^th^ Edition of the Maudsley Prescribing Guidelines^22^.

Based on previously validated methods^12,13^, we adopted a data cut-off of six months before the index date (‘antecedent date’; T-6 months – see Figure 1). This cut-off was introduced to reduce the potential overlap of prodromal symptoms with actual presenting symptoms of FEP as diagnoses are often delayed.

**Figure 1.**
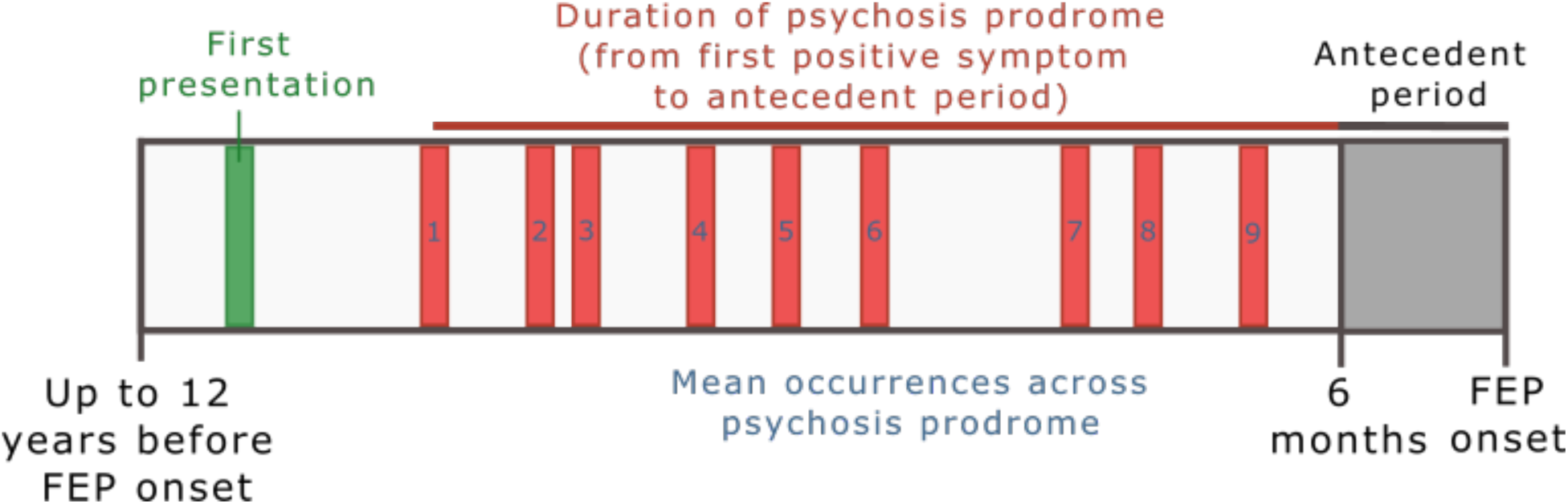
Study design for main analyses. Due to the likely overlap of presenting symptoms in the six months prior to first episode psychosis (FEP) onset, this antecedent period was excluded from analyses. First presentation was considered as the first record of any prodromal feature (not necessarily positive symptom) in the electronic health records. The duration of the psychosis prodrome was defined as the time between the first positive psychotic symptom until the start of the antecedent period. This duration varied between individuals with the maximum duration running between up to 12 years and 6 months before FEP onset. Each individual may have their first occurrence of prodromal features at a different time-point, which leads to varying durations of the duration of the psychosis prodrome. Finally, we compared the mean occurrences of each prodromal feature across the entire prodromal period.

DET+ were defined as patients with FEP who had an accepted referral from OASIS prior to disorder onset. Contrasting with this, DET-were defined as patients with FEP who did not have accepted referral from OASIS prior to disorder onset. We excluded all individuals with data exclusively recorded after the index date or during the antecedent period.

The psychosis prodromal period (measured in months) was defined as the time from the first incidence of any positive psychotic symptom (up to 12 years before index date) until the antecedent date. The overall prodromal period was defined as the time from the first incidence of any of the broadly-defined prodromal features (see definition below) until the antecedent date (Figure 1).

### Variables

At the index date, data was extracted from structured text on age, gender, self-reported ethnicity^23^ (eTable 3), prescribed medication (antidepressants, antipsychotics and mood stabilisers), and ICD-10 diagnoses (eTable 2).

Throughout the prodromal period, NLP algorithms were utilised to extract monthly structured data on the presence or absence (i.e. binary) of symptoms and substance use within each month from unstructured EHR information (eMethods 1, eTable 4). These features were all initiated during the prodromal period and could have either resolved prior to or continued beyond FEP onset^24^. Ultimately, 65 NLP-derived prodromal features with precision ≥80% (mean= 89%) were extracted and stratified into eight previously validated prodromal clusters (eTable 5): catatonic symptoms, depressive symptoms, disorganised symptoms, manic symptoms, negative symptoms, positive symptoms, substance use and other symptoms. This categorization, developed by Jackson et al. (14), is based on previous studies that utilised symptomatology factor analysis^25,26^ and is aligned with publicly available, validated NLP dictionaries^27^. This maximises reliability while simultaneously preserving real-world clinical interpretability and facilitates large-scale clinical pattern identification, crucial for evaluating treatment effectiveness and characterizing interventions, symptom profiles, and outcome-influencing factors. Therefore, each of the eight prodromal clusters is pragmatically relevant for clinical decisions in the context of secondary mental healthcare. However, as shown in eTable 5, these eight prodromal clusters are not completely independent because a few of the prodromal features (e.g. weight loss, apathy, and visual hallucinations) are included in multiple prodromal clusters. This overlap represents transdiagnostic phenomena spanning across a range of clinical dimensions as they are observed in real-world clinical practice. Finally, to fully analyse the independent impact of each individual prodromal feature we also conducted a more fine-grained analysis employing prodromal features as opposed to broader prodromal clusters. When multiple first prodromal features/clusters were recorded during the same month, these were considered to have occurred simultaneously.

### Statistical Analysis

Initial analyses compared differences in positive psychotic symptoms between DET+ and DET-groups before we compared any prodromal features in a subsequent step. First, the proportion of DET+ and DET-individuals who experienced a psychosis prodrome (at least one positive psychotic symptom during the prodromal period) was compared through a logistic regression, reporting Odds Ratios (OR) with 95% confidence intervals (95% CIs).

Second, the duration of the psychosis prodrome (duration of time between first instance of a positive psychotic symptom and the start of the antecedent period) and the overall prodrome (duration of time between first instance of any feature and the start of the antecedent period) was compared between DET+ and DET-individuals. Assumptions for independent t-test were assessed using Shapiro’s test for normality^28^ and Levene’s test for homogeneity of variance^29^. If assumptions were met, groups were compared using independent t-test and effect size estimates were generated using Cohen’s d (d)^30^ and 95% CIs. If assumptions were not met, groups were compared using Mann Whitney U test^31^, Wilcoxon effect size (r)^32^, and 95% CIs.

Third, the incidence of prodromal clusters and features at first presentation (i.e., start of the overall prodrome) were compared between DET+ and DET-individuals through logistic regression, reporting ORs with corresponding 95% CIs.

Fourth, we compared the mean occurrences of prodromal clusters and features across the prodromal period between DET+ and DET-individuals using the same statistical methods as comparing prodromal duration.

Corrections for multiple comparisons were performed using Benjamini-Hochberg procedure with false discovery rate set at 5%. Effect sizes were appraised using pre-defined thresholds: d<0.2/r<0.1 “negligible”, 0.2≤d<0.5/0.1≤r<0.3 “small”, 0.5≤d<0.8/0.3≤r<0.5 “medium”, and d≥0.8/r≥0.5 “large”. The level of significance was set as p<.05 when frequentist statistics were conducted.

### Sensitivity Analyses

We repeated analyses for all outcomes using i) a propensity score-matched sample (to ensure results were not explained by sociodemographic or clinical variables) and ii) a sample with a 1-month antecedent period (to capture a larger proportion of the underlying FEP sample at the expense of potentially capturing more FEP presenting symptoms). Propensity score matching was conducted matching groups on age, gender, ethnicity, psychosis type (affective, non-affective or substance-induced), and medication (antidepressants, antipsychotics and mood stabilisers) at index date using logistic regression and nearest neighbour matching.

All analyses were conducted in R version 4.2.3 employing the effectsize (version 0.8.1) and rstatix (version 0.7.2) packages.

## Results

The analytic sample consisted of 1,545 FEP individuals (31.7% of the overall FEP population with sufficient follow-back data). Of these, 119 had accepted referrals by OASIS (7.7%; DET+) and the remaining 1,426 FEP were undetected (92.3%; DET-) (Table 1).

**Table 1.** Sociodemographic variables at time of index psychotic diagnosis stratified by individuals detected (DET+) and undetected (DET-) by CHR-P services.

|  | Total sample<br>n=1,545 | DET+<br>n=119 | DET-<br>n=1,426 | p-value |
| --- | --- | --- | --- | --- |
| **Age (mean, SD)** | 28.7 (9.4) | 24.7 (5.1) | 29.1 (9.6) | <0.001 |
| **Gender (% Female)** | 593 (38.4%) | 41 (34.5%) | 552 (38.7%) | 0.40 |
| **Ethnicity** |  |  |  | 0.20 |
| Asian | 115 (7.4%) | 16 (13.4%) | 99 (6.9%) |  |
| Black | 733 (47.4%) | 53 (44.5%) | 680 (47.7%) |  |
| Mixed | 67 (4.3%) | 4 (3.4%) | 63 (4.4%) |  |
| White | 454 (29.4%) | 31 (26.1%) | 423 (29.7%) |  |
| Other | 106 (6.9%) | 10 (8.4%) | 96 (6.7%) |  |
| Missing | 70 (4.5%) | 5 (4.2%) | 65 (4.6%) |  |
| **Psychosis type** |  |  |  | 0.30 |
| Affective | 218 (14.1%) | 11 (9.2%) | 207 (14.5%) |  |
| Non-affective | 1,262 (81.6%) | 102 (85.7%) | 1,160 (81.3%) |  |
| Substance-induced | 65 (4.2%) | 6 (5.0%) | 59 (4.1%) |  |
| **Antidepressants** | 450 (29.1%) | 39 (32.8%) | 411 (28.8%) | 0.40 |
| **Antipsychotics** | 1,182 (76.5%) | 77 (64.7%) | 1,105 (77.5%) | 0.041 |
| **Mood stabilisers** | 129 (8.3%) | 4 (3.4%) | 125 (8.8%) | 0.002 |

### Presence of a psychosis prodrome

85.0% of DET+ and 85.6% of DET-individuals endorsed a positive psychotic symptom over the course of the overall prodromal period. There was no significant difference between groups (OR=0.95, 95%CIs=0.58-1.64, p=0.83).

### Duration of the prodrome

The mean duration of the psychosis prodrome was 18.8 months in DET+. (SD=19.3, median [IQR]=13 [2.7]) and 18.4 months in DET-(SD=17.5, median [IQR]=12 [22]) (Figure 2A). This difference was not statistically significant (r=0.004, 95%CI=0.001-0.06, p=0.89).

**Figure 2.**
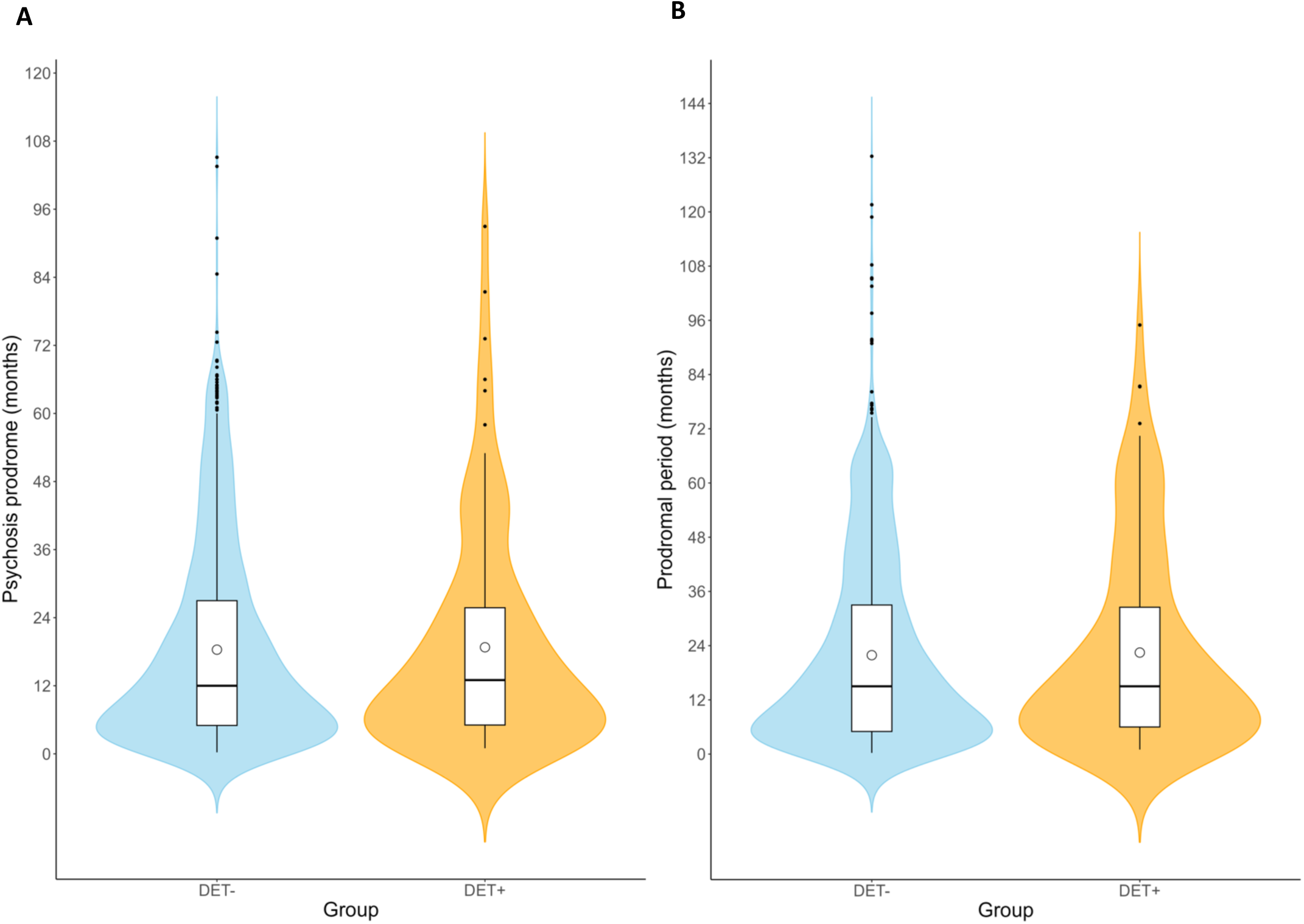
Duration of the A: psychosis prodrome; B: prodromal period. Violin plots representing the distribution of the duration of the prodromal period (months). The circle shape in the box plots represents mean values, the lower end of the box represents the lower quartile, the upper end of the box represents the upper quartile and the horizontal bar median values; vertical lines on each side of the box indicate the maximum and minimum values with the black points representing outliers.

The mean duration of the overall prodrome was 22.5 months in DET+ (SD=22.0, median [IQR]=15 [26.5]) and 21.9 months in DET-individuals (SD=20.7, median [IQR]=15 [28] months) (Figure 2B). This difference was not statistically significant (r=0.002, 95%CI=0.001-0.06, p=0.93).

### First presentation of prodromal clusters and features

There was no significant difference in the proportion of DET+ individuals who exhibited any positive psychotic symptoms at first presentation (68%) compared to DET-individuals (69%; OR=0.96; 95%CIs=0.56-1.66) (Figure 3A).

**Figure 3.**
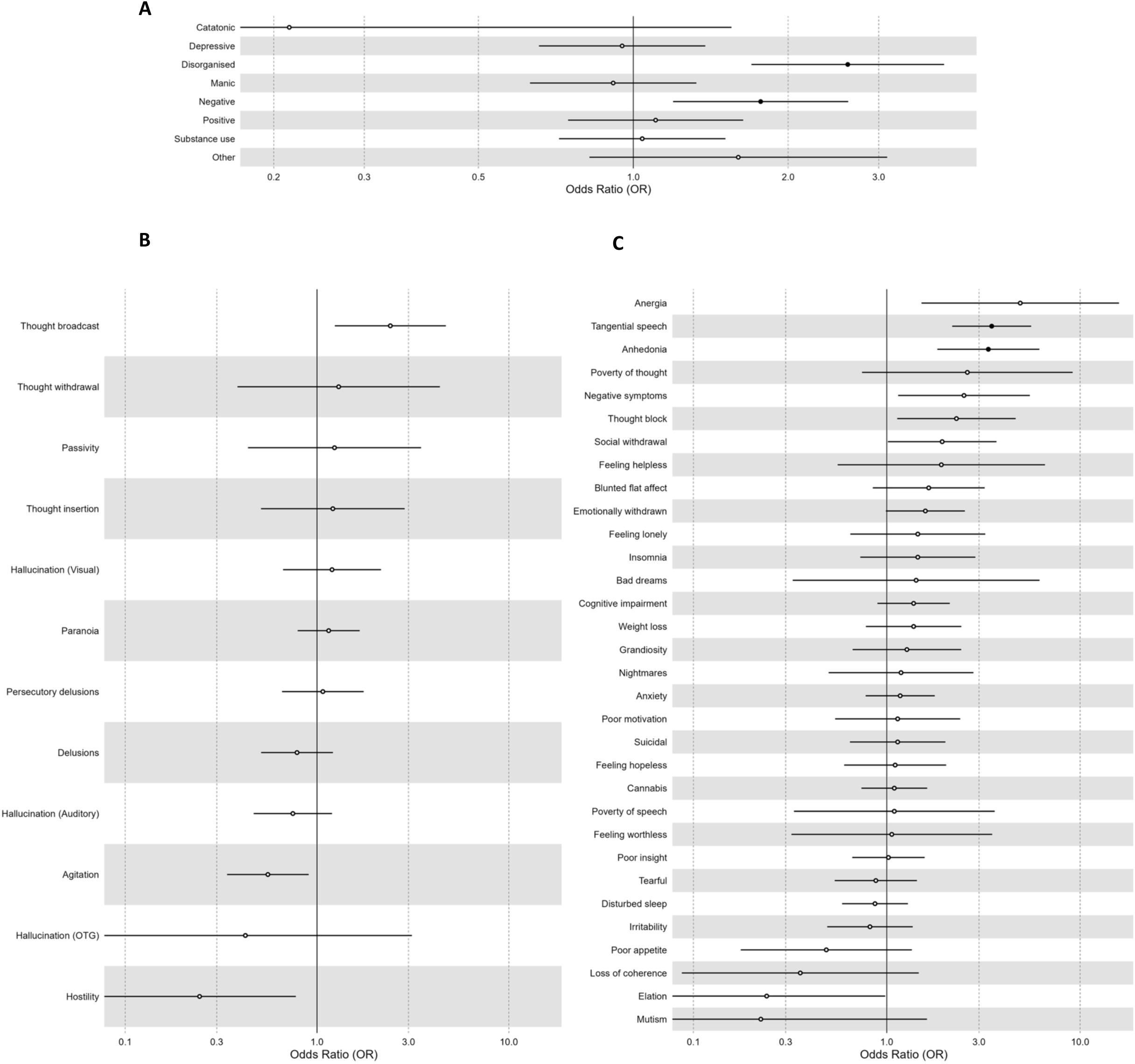
Forest plot comparing first presentation of prodromal A clusters; B psychotic symptoms; and C other symptoms and substance use across DET+ and DET-individuals. OR>1 indicates higher prevalence of the cluster/feature in DET+ individuals compared to DET-. OR<1 indicates lower prevalence of the cluster/feature in DET+ individuals compared to DET-. Dark circles indicate clusters/features that were significantly different from 0 following Benjamini-Hochberg correction. *Abbreviations: OTG, olfactory, tactile or gustatory*.

There were no differences in proportion of DET+ and DET-individuals who endorsed individual psychotic symptoms at first presentation (Figure 3B).

DET+ showed a higher incidence within the disorganised symptom cluster (DET+=0.29; DET- =0.13; OR=2.60; 95%CIs=1.68-4.20, p_corr_<0.001), as well as of anhedonia (DET+=0.13; DET- =0.04; OR=3.34; 95%CIs=1.77-6.20, p_corr_<0.001) and tangential speech (DET+=0.22; DET-=0.07; OR=3.48; 95%CIs=2.13-6.20, p_corr_<0.001) compared to DET- (Figure 3C).

All other clusters and features displayed non-significant group differences (p_corr_>0.05; Figure 3A-C; eTable 7).

### Occurrences of clusters and features across the prodromal period

There were no significant differences in the occurrences of positive symptoms across the prodromal period (DET+=4.82 [SD=5.86]; DET-=4.30 [SD=5.25]; r=0.03; 95%CIs=0.001-0.08; p_corr_=0.56) (Figure 4A). DET+ showed a higher number of occurrences of thought broadcasting MD=0.13, r=0.07, 95%CIs=0.01-0.14, p_corr_=0.04) and lower number of occurrences of hostility (MD=-0.26, r=0.08, 95%CIs=0.03-0.11, p_corr_=0.04) compared to DET-, though effect sizes were small (Figure 4B).

**Figure 4.**
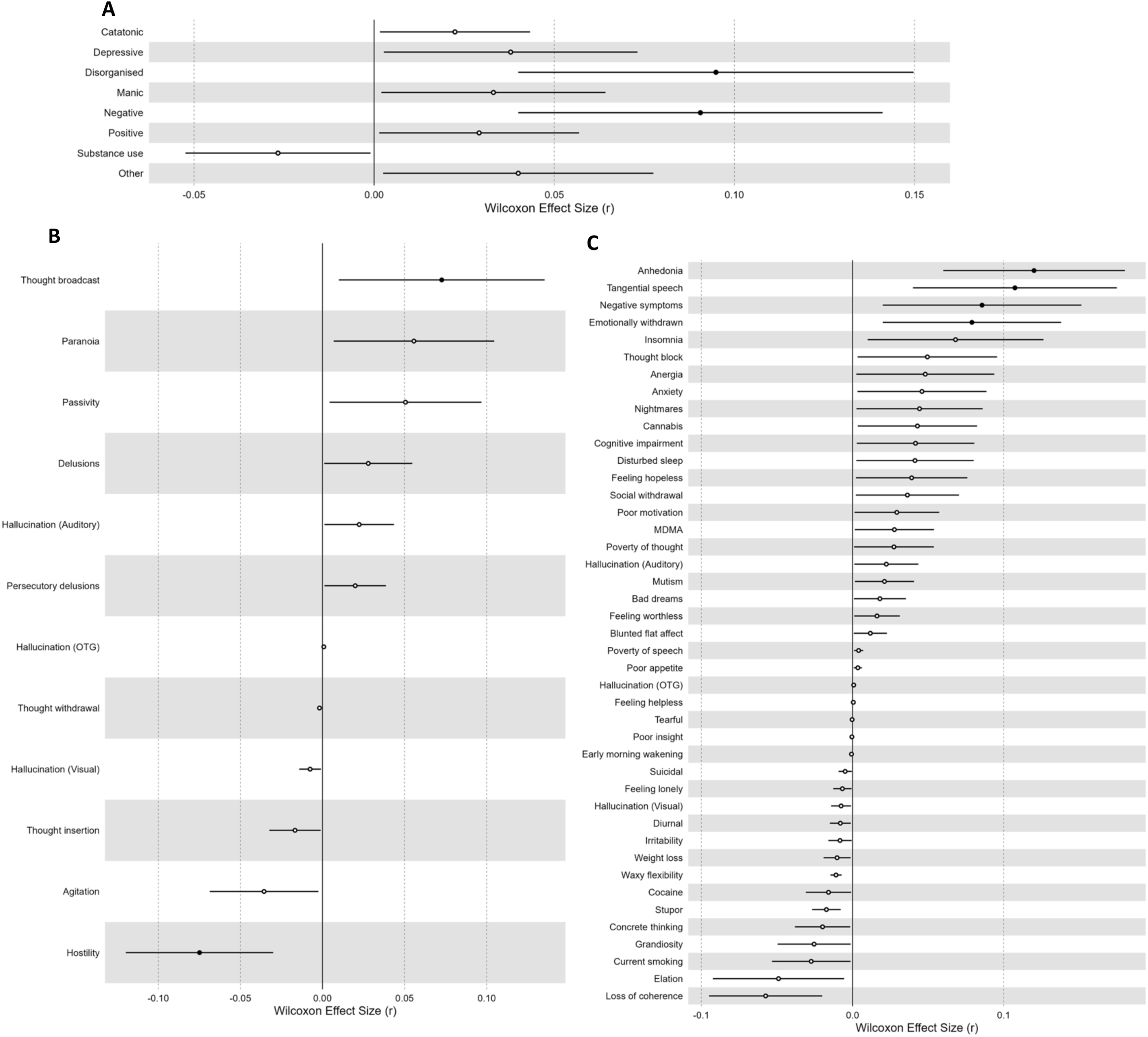
Forest plot of difference in number of occurrences of prodromal A clusters; B psychotic symptoms; and C other symptoms and substance use across the overall prodromal periods between DET+ and DET-individuals. r>0 indicates higher prevalence of the cluster/feature in DET+ individuals compared to DET-. r<0 indicates lower prevalence of the cluster/feature in DET+ individuals compared to DET-. Dark circles indicate clusters/features that were significantly different from 0 following Benjamini-Hochberg correction. *Abbreviations: OTG, olfactory, tactile or gustatory*.

DET+ showed a higher number of occurrences in disorganised (MD=0.41, r=0.10, 95%CIs=0.04-0.15, p_corr_=0.006), and negative (MD=0.76, r=0.09, 95%CIs=0.04-0.14, p_corr_=0.009) symptom clusters and tangential speech (MD=0.35, r=0.11, 95%CIs=0.04-0.17, p_corr_=0.001), anhedonia (MD=0.35, r=0.12, 95%CIs=0.06-0.18, p_corr_<0.001), negative symptoms (MD=0.76, r=0.09, 95%CIs=0.02-0.15, p_corr_=0.01) and emotional withdrawal (MD=0.27, r=0.08, 95%CIs=0.02-0.14, p_corr_=0.03) than in DET-, though effect sizes were negligible/small (Figure 4C).

All other clusters and features displayed non-significant group differences (p_corr_>0.05; Figure 4A-C; eTable 8).

### Sensitivity Analyses: Propensity Score Matched Sample

Following propensity score matching, a final sample of 238 individuals (119 DET+ and 119 matched DET-) were included in sensitivity analyses. The propensity matched DET- sample was similar in terms of demographic and clinical features to the DET+ sample but displayed differences from the overall DET-sample (eTable 6).

There were no significant differences in duration of the psychosis prodrome (r=0.02, 95%CIs=0.002-0.16, p=0.73) or the prodromal period (r=0.02, 95%CI=0.002-0.15, p=0.71) between the final DET+ and DET-samples.

For first presentation, only the incidence of disorganised symptom cluster (DET+=0.29; DET- =0.12; OR=3.00; 95%CIs=1.54-6.20, p_corr_<0.001) and tangential speech feature (DET+=0.22; DET-=0.07; OR=3.88; 95%CIs=1.75-10.20, p_corr_<0.001) remained significantly higher in DET+ compared to DET- (eTable 9).

For occurrences across the prodrome, all effects were negligible/small (eTable 10).

### Sensitivity Analyses: 1-month Antecedent Period

The shorter antecedent period expanded our sample to 2,787 FEP individuals (55.6% of the overall FEP population). 76.4% of DET+ and 69.8% of DET-individuals endorsed a positive psychotic symptom over the course of the overall prodromal period. There was no significant difference between groups (OR=1.39, 95%CIs=0.9-2.23, p=0.15).

The duration of the psychosis prodrome was shorter in DET-than DET+ but the effect size was negligible (r=0.02, 95%CIs=0.004-0.04, p=0.04). There was no significant difference in duration of the prodromal period (r=0.02, 95%CIs=0.002-0.03, p=0.06).

For first presentation, DET+ individuals presented with agitation (DET+=0.20; DET-=0.36; OR=0.45; 95%CIs=0.3-0.64, p_corr_<0.001), delusions (DET+=0.28; DET-=0.39; OR=0.60; 95%CIs=0.42 - 0.83, p_corr_=0.02) and hostility (DET+=0.06; DET-=0.14; OR=0.41; 95%CIs=0.21 - 0.72, p_corr_=0.03) less frequently compared to DET-(eTable 11).

The incidence of the negative symptom cluster remained higher in DET+ compared to DET-but disorganised symptoms were no longer significant. Anhedonia and tangential speech remained significantly higher in DET+. DET+ individuals presented with emotional withdrawal (DET+=0.26; DET-=0.16; OR=1.83; 95%CIs=1.28-2.58, p_corr_=0.01), social withdrawal (DET+=0.12; DET-=0.06; OR=2.10; 95%CIs=1.28-3.31, p_corr_=0.02) and anergia (DET+=0.03; DET- =0.01; OR=3.94; 95%CIs=1.29-9.93, p_corr_=0.04) more frequently and elation (DET+=0.04; DET- =0.11; OR=0.34; 95%CIs=0.14-0.67, p_corr_=0.03) less frequently compared to DET- (eTable 11). For occurrences across the prodrome, all effects were negligible/small (eTable 12).

## Discussion

To our knowledge, this is the first study utilising NLP and EHR data to compare the FEP prodrome of individuals detected (DET+) and undetected (DET-) by CHR-P services. The key finding was that the prodromal phase of DET+ and DET-are very similar, with only negligible differences in first presentation and occurrences of prodromal features and clusters across the prodromal period. Together, this evidence suggests that psychotic symptom-focused detection strategies targeting DET+ can be equally successful in DET-.

There is a detectable psychosis prodrome to FEP. In line with our previous analyses, using less stringent definitions of psychosis^13^, the duration of this prodrome was found to be close to 2 years (∼18 months), with no difference between DET+ and DET-individuals. This represents a clear targetable period for detection that is in line with the duration of care offered by CHR-P services^33,34^. Generally, however, the psychosis prodrome is considered to be even longer than this, extending up to 138 months^25^, suggesting that our sample may have experienced delays in help-seeking or accessing care following the first onset of symptoms/features^35^. This may also reflect that an individual’s symptoms would need to be relatively severe to access secondary care and therefore would only be represented in our dataset following substantial symptomatic exacerbation.

This prodrome includes positive psychotic symptoms with around 85% of both DET+ and DET-individuals endorsing at least one positive psychotic symptom over the course of the prodrome. While we were unable to measure severity and frequency, as required for psychometric CHR-P assessments, these findings are suggestive of a CHR-P state prior to FEP onset and are similar to meta-analytic estimates (78.3%).^8^

We similarly found limited differences in how DET+ and DET-individuals first present to secondary care and their features across the prodrome, both in terms of positive psychotic symptoms but also in terms of other psychiatric symptoms and substance use. 92% of prodromal features did not differ between DET+ and DET-either at first presentation or across the prodrome. However, DET+ individuals were more likely to present with disorganised symptoms, particularly tangential speech, and displayed higher occurrences of disorganised and negative symptom clusters across the prodrome, namely anergia and anhedonia. These disorganised symptoms may result in concerns from friends and family that may lead to earlier referrals and help-seeking. Although positive psychotic features (e.g., delusions) may be more expected in CHR-P individuals^36,37^, negative and disorganised symptoms are also common in CHR-P individuals^38^ and are strongly associated with both each other^39^ and transition to psychosis^40,41^. Negative symptoms generally develop before positive symptoms^42,43^ which may lead to early help seeking behaviour e.g. to GPs^44^ or family/friends^36,37,45^. All other prodromal clusters and features were no different between DET+ and DET-subgroups, and only disorganised symptoms and tangential speech remained significantly more prevalent at first presentation in the DET-propensity matched sample despite substantial differences in case-mix with the overall DET-sample.

Together, this evidence suggests that the psychosis prodromes experienced by DET+ and DET-individuals are not substantially different. The lack of differences suggest that the psychometric assessments used to detect CHR-P individuals are appropriate and are able to capture relevant prodromal symptoms in DET-individuals. Instead, the low proportion of DET+ cases is likely caused by inefficient detection strategies failing to identify the right people to assess, both at the service- and individual-levels. Improving current detection strategies can therefore have substantial impact on improving the CHR-P paradigm^7,46^. Paradoxically, however, improving outreach can improve detection efforts but is likely to dilute pre-test risk. Expanding outreach efforts can lead to recruiting from a wider population with a lower risk of psychosis. For example, the incidence of psychosis within secondary mental healthcare is substantially higher than in the general population. Expanding outreach to settings with lower inherent psychosis risk, like the general population, will lead to a lower psychosis risk within the population you are screening i.e. lower pre-test risk. Due to the prognostic accuracy of psychometric assessments for CHR-P, screening 100,000 people in the general population will lead to roughly 99,999 false positives, compared to 82,640 when screening 100,000 people in secondary care,^7^ therefore reducing the underlying risk of CHR-P samples^47,48^. Any efforts to improve detection, must rely on targeted approaches that are able to enrich risk.

Therefore, complementary strategies have been proposed to improve detection of CHR-P individuals using digital health data targeting the community, primary care and secondary care. Firstly, self-reported pre-screening instruments, like the prodromal questionnaire (PQ-16), can be used online to identify probable CHR-P cases in the community at a similar standard to typical referral pathways^49^, without diluting risk^7^. This can potentially be refined further by combining this with other assessments of psychosis risk, such the Psychosis Polyrisk Score^50–52^, a multivariate assessment of environmental risk, which is being tested in an ongoing study (E-

Detection Tool for Emerging Mental Disorders; ENTER), or longitudinal measurement of symptomatology through digital phenotyping methods^53,54^. Secondly, P-Risk is an EHR-based clinical prediction model for psychosis developed, internally^55^ and externally validated^56^ in primary care data, displaying robust performance (C=0.79). Previous research has suggested that primary care clinicians are not confident in ascertaining psychosis risk so P-Risk can serve a key role in improving referrals from primary care. Finally, a transdiagnostic risk calculator for psychosis in secondary care EHRs has been developed^4^, extensively externally validated (C=0.68-0.79^57–60)^ and implemented in real world clinical practice^61,62^ (summarised in ^63^). Refined versions of the risk calculator have since been tested showing improved performance (C=0.85-0.89)^64,65^. Together, these strategies can improve our ability to detect CHR-P individuals at scale and extend the benefits of care to more people prior to FEP onset.

Moreover, more than half of our sample first presented with depressive, manic and/or other symptoms, in addition to positive psychotic symptoms, highlighting the transdiagnostic prodromal characteristics experienced prior to FEP. Acknowledging the overlapping prodromal features across mental disorders^12,66,67^, transdiagnostic early detection services^68–73^ could be an alternate route forward to effective preventive psychiatry^74^.

This study has some limitations. Firstly, although EHRs have high ecological validity^75^, the features recorded in clinical notes were not psychometrically validated. Furthermore, while there is evidence that data recorded in EHRs are usually predictive of true validated diagnoses^75^, the use of structured diagnostic interviews are susceptible to selection bias, particularly as NLP tools create noise in terms of clinician subjectivity, including structural or unconscious biases, which may influence how symptoms are recorded^76^, thus impacting standardisation. However, this was partially mitigated by use of NLP algorithms which have a level of ≥80% precision. Secondly, our estimates are representative of detectable prodromes and prodromal features recorded in secondary care, while the true prodrome is likely longer, with prodromal symptoms starting prior to people seeking help and accessing secondary care, and more common, in line with previous research^8,35^. Similarly, we were only able to assess the prodromes in individuals with sufficient follow-back present in the EHR; fortunately, those included in our sample appear to be similar to those excluded, including in sensitivity analyses reducing the duration of our antecedent period. Future research will require densely phenotyped longitudinal cohorts with regular symptomatological assessments, which could be facilitated by digital health methods^8,77,78^. Thirdly, there were some differences in clinical features between the two groups, which could exacerbate differences in prodromal features but these were negligible in the main analyses and consistent with our propensity score matched cohort. Finally, we were not able to externally validate our findings, so future work is needed to test whether these results generalise to other settings without CHR-P services or a lower incidence of psychosis.

In conclusion, DET+ and DET-individuals experience similar prodromes prior to FEP onset, including positive symptoms, other psychiatric symptoms and substance use. DET-individuals can likely be identified earlier if detection strategies are improved.

## Data Sharing Statement

Data are owned by a third party, Maudsley Biomedical Research Centre (BRC) Clinical Records Interactive Search (CRIS) tool, which provides access to anonymised data derived from SLaM electronic medical records. These data can only be accessed by permitted individuals from within a secure firewall (i.e. the data cannot be sent elsewhere), in the same manner as the authors. For more information please contact.

All analysis code is available on GitHub: https://github.com/dapoliver/FEP_Prodrome.

## Ethics committee approval

Permissions for the study were granted by the Oxfordshire Research Ethics Committee C (23/SC/0257); because the data set comprised deidentified data, informed consent was not required^79^.

## Authors’ contribution

DO designed the study. MI, MG, MA and DO ran the statistical analyses. MI, MG and DO drafted the manuscript. All authors contributed to critical revision of the manuscript for important intellectual content and approved the final version of the manuscript.

## Data Availability

The data accessed by CRIS remain within an NHS firewall and governance is provided by a patient-led oversight committee. Subject to these conditions, data access is encouraged and those interested should contact Robert Stewart, CRIS academic lead. Further details regarding the CRIS platform can be found elsewhere. All analysis code is available on GitHub: https://github.com/dapoliver/ProdromeFEP.

