## Supplementary Material for "Can we detect the undetected? Comparing the prodromes of individuals with first episode psychosis detected and undetected by clinical high risk for psychosis services: an electronic health record study"

Inns M, Gill M, Arribas M et al. Characterising the duration and first presentation of the psychosis prodrome in individuals detected and undetected by at-risk mental state services: an electronic health record study.

### Table of Contents

**Methods**

eTable 1: The RECORD statement

eTable 2: Operationalisation of ICD-10 diagnoses.

eFigure 1 Study design.

eTable 3: Definition of self-reported ethnicity

eMethods 1: NLP algorithm development and validation

eTable 4: Type and precision for 65 NLP algorithms

eTable 5: Prodromal clusters utilised to stratify 65 prodromal features investigated in this research.

**Results**

eTable 6: Baseline sociodemographic variables in overall and propensity score matched samples.

eTable 7: Results table for first presentation

eTable 8: Results table across prodrome

eTable 9: Results table for first presentation in Propensity Score Matched Sample

eTable 10: Results table across prodrome in Propensity Score Matched Sample

eTable 11: Results table for first presentation using 1-month antecedent period sample

eTable 12: Results table across prodrome using 1-month antecedent period sample

eReferences

Methods

**eTable 1** The RECORD statement – checklist of items, extended from the STROBE statement, that should be reported in observational studies using routinely collected health data.

| | Item No. | STROBE items | Location in manuscript where items are reported | RECORD items | Location in manuscript where items are reported |
|---|---|---|---|---|---|
| **Title and abstract** | | | | | |
| | 1 | (a) Indicate the study's design with a commonly used term in the title or the abstract (b) Provide in the abstract an informative and balanced summary of what was done and what was found | Abstract | RECORD 1.1: The type of data used should be specified in the title or abstract. When possible, the name of the databases used should be included.<br><br>RECORD 1.2: If applicable, the geographic region and timeframe within which the study took place should be reported in the title or abstract. | Abstract<br><br>Abstract |

| | | | | | |
|---|---|---|---|---|---|
| | | | | RECORD 1.3: If linkage between databases was conducted for the study, this should be clearly stated in the title or abstract. | NA |
| Introduction | | | | | |
| Background rationale | 2 | Explain the scientific background and rationale for the investigation being reported | Introduction | | |
| Objectives | 3 | State specific objectives, including any prespecified hypotheses | Abstract, Introduction, and Methods | | |
| Methods | | | | | |
| Study Design | 4 | Present key elements of study design early in the paper | Abstract and Methods | | |
| Setting | 5 | Describe the setting, locations, and relevant dates, including periods of recruitment, exposure, follow-up, and data collection | Abstract and Methods | | |

| | | | | | |
|---|---|---|---|---|---|
| Participants | 6 | *(a) Cohort study* - Give the eligibility criteria, and the sources and methods of selection of participants. Describe methods of follow-up<br>*Case-control study* - Give the eligibility criteria, and the sources and methods of case ascertainment and control selection. Give the rationale for the choice of cases and controls<br>*Cross-sectional study* - Give the eligibility criteria, and the sources and methods of selection of participants<br><br>*(b) Cohort study* - For matched studies, give matching criteria and number of exposed and unexposed | Abstract and Methods<br><br>NA<br><br>NA<br><br>NA<br><br>Methods | RECORD 6.1: The methods of study population selection (such as codes or algorithms used to identify subjects) should be listed in detail. If this is not possible, an explanation should be provided.<br><br>RECORD 6.2: Any validation studies of the codes or algorithms used to select the population should be referenced. If validation was conducted for this study and not published elsewhere, detailed methods and results should be provided.<br><br>RECORD 6.3: If the study involved linkage of databases, consider use of a flow diagram or other graphical display to demonstrate the data linkage | Methods<br><br>Methods, supplementary, and referenced previous publications addressing the codes or algorithms used<br><br>NA |

| | | | | | |
|---|---|---|---|---|---|
| | | *Case-control study* - For matched studies, give matching criteria and the number of controls per case | | process, including the number of individuals with linked data at each stage. | |
| Variables | 7 | Clearly define all outcomes, exposures, predictors, potential confounders, and effect modifiers. Give diagnostic criteria, if applicable. | Methods, and supplementary material | RECORD 7.1: A complete list of codes and algorithms used to classify exposures, outcomes, confounders, and effect modifiers should be provided. If these cannot be reported, an explanation should be provided. | Materials and methods, supplementary material |
| Data sources/ measurement | 8 | For each variable of interest, give sources of data and details of methods of assessment (measurement).<br>Describe comparability of assessment methods if there is more than one group | Methods | | |
| Bias | 9 | Describe any efforts to address potential sources of bias | Methods and Discussion | | |

| | | | |
|---|---|---|---|
| Study size | 10 | Explain how the study size was arrived at | Methods and supplementary material |
| Quantitative variables | 11 | Explain how quantitative variables were handled in the analyses. If applicable, describe which groupings were chosen, and why | Statistical analysis |
| Statistical methods | 12 | (a) Describe all statistical methods, including those used to control for confounding<br>(b) Describe any methods used to examine subgroups and interactions<br>(c) Explain how missing data were addressed<br>(d) *Cohort study* - If applicable, explain how loss to follow-up was addressed | Statistical analysis<br>Statistical analysis<br>NA<br>NA<br>NA |

| | | | | | |
|---|---|---|---|---|---|
| | | *Case-control study* - If applicable, explain how matching of cases and controls was addressed<br>*Cross-sectional study* - If applicable, describe analytical methods taking account of sampling strategy<br>(e) Describe any sensitivity analyses | NA<br><br>Statistical analysis | | |
| Data access and cleaning methods | | . | | RECORD 12.1: Authors should describe the extent to which the investigators had access to the database population used to create the study population.<br>RECORD 12.2: Authors should provide information on the data cleaning methods used in the study. | Methods and supplementary material<br><br>NA |
| Linkage | | .. | | RECORD 12.3: State whether the study included person-level, institutional-level, or other data linkage across two or more databases. The methods of | NA |

| | | | | | |
|---|---|---|---|---|---|
| | | | | linkage and methods of linkage quality evaluation should be provided. | |
| Results | | | | | |
| Participants | 13 | (a) Report the numbers of individuals at each stage of the study (*e.g.*, numbers potentially eligible, examined for eligibility, confirmed eligible, included in the study, completing follow-up, and analysed)<br>(b) Give reasons for non-participation at each stage.<br>(c) Consider use of a flow diagram | Results<br><br>NA<br><br>NA | RECORD 13.1: Describe in detail the selection of the persons included in the study (*i.e.,* study population selection) including filtering based on data quality, data availability and linkage. The selection of included persons can be described in the text and/or by means of the study flow diagram. | Methods, results, and supplementary material |
| Descriptive data | 14 | (a) Give characteristics of study participants (*e.g.*, demographic, clinical, social) and information on exposures and potential confounders | supplementary material<br><br>NA | | |

| | | | |
|---|---|---|---|
| | | (b) Indicate the number of participants with missing data for each variable of interest<br>(c) *Cohort study* - summarise follow-up time (*e.g.*, average and total amount) | Results |
| Outcome data | 15 | *Cohort study* - Report numbers of outcome events or summary measures over time<br><br>*Case-control study* - Report numbers in each exposure category, or summary measures of exposure<br>*Cross-sectional study* - Report numbers of outcome events or summary measures | Results and supplementary material<br><br>NA<br><br>NA |
| Main results | 16 | (a) Give unadjusted estimates and, if applicable, confounder-adjusted estimates and their | Results and supplementary material |

| | | | |
|---|---|---|---|
| | | precision (e.g., 95% confidence interval). Make clear which confounders were adjusted for and why they were included<br>(b) Report category boundaries when continuous variables were categorized<br>(c) If relevant, consider translating estimates of relative risk into absolute risk for a meaningful time period | <br><br><br><br><br>NA<br><br><br>NA |
| Other analyses | 17 | Report other analyses done—e.g., analyses of subgroups and interactions, and sensitivity analyses | Results |
| Discussion | | | |
| Key results | 18 | Summarise key results with reference to study objectives | Discussion (headed sections significant symptoms, prodrome length |

| | | | | | |
|---|---|---|---|---|---|
| | | | and common features) | | |
| Limitations | 19 | Discuss limitations of the study, taking into account sources of potential bias or imprecision. Discuss both direction and magnitude of any potential bias | Discussion (methodological considerations) | RECORD 19.1: Discuss the implications of using data that were not created or collected to answer the specific research question(s). Include discussion of misclassification bias, unmeasured confounding, missing data, and changing eligibility over time, as they pertain to the study being reported. | Discussion |
| Interpretation | 20 | Give a cautious overall interpretation of results considering objectives, limitations, multiplicity of analyses, results from similar studies, and other relevant evidence | Discussion | | |

| | | | | | |
|---|---|---|---|---|---|
| Generalisability | 21 | Discuss the generalisability (external validity) of the study results | Discussion | | |
| Other Information | | | | | |
| Funding | 22 | Give the source of funding and the role of the funders for the present study and, if applicable, for the original study on which the present article is based | NA | | |
| Accessibility of protocol, raw data, and programming code | | .. | | RECORD 22.1: Authors should provide information on how to access any supplemental information such as the study protocol, raw data, or programming code. | Statistical analysis, supplementary material |

**eTable 2:** Operationalisation of ICD-10 diagnoses utilised in the current research.

| Specific ICD-10 code | Specific ICD-10 diagnosis, version 2016 | Psychosis Type |
|---|---|---|
| [F10-F19].4, [F10-F19].5, [F10-F19].7 | Mental and behavioural disorders due to psychoactive substance use with psychotic symptoms or delirium | Substance Induced |
| F20-29 | Schizophrenia, schizotypal and delusional disorders | Non-affective |
| F30.2 | Mania with psychotic symptoms | Affective |
| F31.2 | Bipolar affective disorder, current episode manic with psychotic symptoms | Affective |
| F31.5 | Bipolar affective disorder, current episode severe depression with psychotic symptoms | Affective |
| F32.3 | Severe depressive episode with psychotic symptoms | Affective |
| F33.3 | Recurrent depressive disorder, current episode severe with psychotic symptoms | Affective |
| F53.1 | Severe mental and behavioural disorders associated with the puerperium, not elsewhere classified (post-partum psychosis) | Postpartum |

**eTable 3:** Definition of self-reported ethnicity according to UK Office of National Statistics[2]

| **Ethnic group** | **Self-reported ethnicity as recorded in EHR** |
|---|---|
| Black | Black or Black British - African<br>Black or Black British - Caribbean<br>Black or Black British - Any other Black background |
| White | White - British<br>White - Irish<br>White - Any other White background |
| Asian | Asian or Asian British - Bangladeshi<br>Asian or Asian British - Indian<br>Asian or Asian British - Pakistani<br>Asian or Asian British - Any other Asian background<br>Other Ethnic Groups - Chinese |
| Mixed | Mixed - White and Asian<br>Mixed - White and Black African<br>Mixed - White and Black Caribbean<br>Mixed - Any other mixed background |
| Other | Other Ethnic Groups - Any other ethnic group |
| Missing | Not Known<br>Not Recorded |

**eMethods 1:** NLP algorithm development and validation

CRIS algorithms have been developed with machine learning approaches against gold standard training sets manually annotated for positive, negative, and unknown or irrelevant mentions, therefore they are able to exclude language features including negation (e.g., patient denies guilt) and irrelevant mentions (e.g., his father felt guilty). Failure patterns driving false positives were identified through manual testing of algorithm output (e.g., "*name* was found guilty of stealing"). The machine learning classifier trained on these false positives to ignore these and similar statements in an iterative process of testing and redeveloping until acceptable precision was achieved. Patterns of failure identified through testing can be found in the CRIS service's comprehensive online NLP algorithm library provided at https://www.maudsleybrc.nihr.ac.uk/facilities/clinical-record-interactive-search-cris/cris-natural-language-processing/ [3].

NLP algorithm performance was measured with precision (proportion of true positive instances of total NLP-labelled positive instances) and recall (proportion of true positive instances of all positive instances in the text). As EHRs grant many opportunities for term detection, precision was favoured over recall, using only NLP algorithms with at least 80% precision (see eTable 4 for a list of NLP algorithms utilised in this study).

Algorithms were manually validated by an independent researcher at the SLaM Biomedical Research Centre Nucleus prior to the current research project. The programme for algorithms validation was responsive to the specific needs of scheduled CRIS research activities and therefore the approach was not standardised.

**eTable 4**: Type and precision for 52 NLP algorithms (adapted from Arribas M, Oliver D, Patel R et al.[1])

Tables includes precision values from the CRIS Natural Language Processing Library (2021)[2], which were identified by randomly selecting positive annotations from each algorithm for a specified cohort, limited to one annotation per patient ID. Precision scores were thereafter calculated as the ratio of the number of relevant (true positive) instances retrieved from the total NLP-labelled positive instances (including irrelevant [false positive] and relevant [true positive] instances) for each NLP algorithm.

| NLP algorithms | Cohort | Annotations validated (n) | Precision (%) |
|---|---|---|---|
| Aggression | Random sample | 100 | 90 |
| Agitation | Random sample | 100 | 85 |
| Anergia | Random sample | 100 | 84 |
| Anhedonia | Random sample | 100 | 94 |
| Anxiety | Random sample | 100 | 94 |
| Apathy | Random sample | 100 | 94 |
| Arousal | Random sample | 100 | 89 |
| Bad dreams | CAMHS events | 100 | 92 |
| Blunted affect | Random sample | 100 | 98 |
| Cannabis use | All patients | 100 | 88 |
| Circumstantiality | Random sample | 100 | 97 |
| Cocaine use | Random sample | 30 | 97 |
| Cognitive impairment | Patients with F20 | 100 | 84 |
| Concrete thinking | Random sample | 146 | 91 |
| Delusional thinking | Random sample | 100 | 90 |
| Derailment | Random sample | 100 | 87 |
| Disturbed sleep | Random sample | 100 | 89 |
| Diurnal mood | Random sample | 100 | 86 |
| Early morning wakening | Random sample | 100 | 96 |
| Echolalia | Random sample | 100 | 96 |
| Elation | Random sample | 100 | 95 |
| Emotional withdrawal | Random sample | 100 | 87 |
| Feeling helpless | Random sample | 100 | 92 |
| Feeling hopeless | Random sample | 100 | 88 |
| Feeling lonely | Random sample | 100 | 87 |
| Feeling worthless | Random sample | 100 | 91 |
| Flight of ideas | Random sample | 100 | 89 |

| | | | |
|---|---|---|---|
| Formal thought disorder | Random sample | 100 | 85 |
| Grandiosity | Random sample | 100 | 89 |
| Guilt | Random sample | 100 | 84 |
| Hallucinations (all) | Random sample | 100 | 90 |
| Hallucinations (auditory) | Random sample | 100 | 92 |
| Hallucinations (OTG: olfactory, tactile, gustatory) | Random sample | 100 | 86 |
| Hallucinations (visual) | Random sample | 100 | 83 |
| Hostility | Random sample | 100 | 86 |
| Insomnia | Random sample | 100 | 97 |
| Irritability | Random sample | 100 | 99 |
| Loss of coherence | Random sample | 158 | 85 |
| Low energy | CAMHS events | 100 | 89 |
| MDMA use | Random sample | 100 | 94 |
| Mood instability | Random sample | 100 | 91 |
| Mutism | Random sample | 100 | 95 |
| Negative symptoms | Random sample | 100 | 87 |
| Nightmares | Random sample | 100 | 89 |
| Paranoia | Random sample | 100 | 89 |
| Passivity | Random sample | 100 | 88 |
| Persecutory ideation | Random sample | 100 | 80 |
| Poor appetite | Random sample | 100 | 89 |
| Poor concentration | Random sample | 100 | 88 |
| Poor insight | Random sample | 100 | 85 |
| Poor motivation | Random sample | 100 | 95 |
| Poverty of speech | Random sample | 100 | 88 |
| Poverty of thought | Random sample | 100 | 98 |
| Social withdrawal | Random sample | 100 | 98 |
| Stupor | Random sample | 100 | 88 |
| Suicidality | CAMHS events | 100 | 87 |
| Tangential speech | Random sample | 100 | 90 |
| Tearfulness | Random sample | 100 | 94 |
| Thought block | Random sample | 100 | 92 |
| Thought broadcast | Random sample | 100 | 84 |
| Thought insertion | Random sample | 100 | 84 |
| Thought withdrawal | Random sample | 100 | 84 |
| Tobacco use | Random sample | 118 | 90 |
| Waxy flexibility | Random sample | 100 | 81 |
| Weight loss | Random sample | 100 | 80 |

**eTable 5**: Prodromal clusters utilised to stratify 65 prodromal features investigated in this research[1].

| Prodromal Cluster (number of prodromal features in the prodromal cluster) | | | | | | | |
|---|---|---|---|---|---|---|---|
| **Catatonic (0-4)** | **Depressive (0-21)** | **Disorganised (0-8)** | **Manic (0-8)** | **Negative (0-12)** | **Positive (0-16)** | **Substance use (0-4)** | **Other (0-8)** |
| Echolalia | Anergia | Circumstantiality | Disturbed sleep | Anergia | Aggression | Cannabis use | Anxiety |
| Mutism | Anhedonia | Derailment of speech | Elation | Anhedonia | Agitation | Cocaine use | Bad dreams |
| Stupor | Apathy | Flight of ideas | Grandiosity | Apathy | Arousal | MDMA use | Cognitive impairment |
| Waxy flexibility | Disturbed sleep | Formal thought disorder | Insomnia | Blunted affect | Delusions | Tobacco use | Feeling lonely |
| | Diurnal mood | Loss of coherence | Irritability | Concrete thinking | Hallucinations (all) | | Hallucinations (visual) |
| | Early morning wakening | Poor concentration | Poor appetite | Emotionally withdrawn | Hallucinations (auditory) | | Mood instability |
| | Guilt | Tangential speech | Poor concentration | Low energy | Hallucinations (OTG) | | Nightmares |
| | Feeling helpless | Thought block | Weight loss | Negative symptoms | Hallucinations (visual) | | Poor insight |
| | Feeling hopeless | | | Poor motivation | Hostility | | |
| | Feeling worthless | | | Poverty of speech | Irritability | | |
| | Insomnia | | | Poverty of thought | Paranoia | | |
| | Low energy | | | Social withdrawal | Passivity | | |
| | Poor appetite | | | | Persecutory delusions | | |
| | Poor concentration | | | | Thought broadcast | | |
| | Poor motivation | | | | Thought insertion | | |
| | Poverty of speech | | | | Thought withdrawal | | |
| | Poverty of thought | | | | | | |
| | Social withdrawal | | | | | | |
| | Suicidality | | | | | | |
| | Tearfulness | | | | | | |
| | Weight loss | | | | | | |

**eTable 6** Sociodemographic variables at time of index psychotic diagnosis in overall and propensity score matched sample.

| | Overall (n=1,545) | DET- (n=119)[1] | DET+ (n=119) | Excluded (n=3,467) |
|---|---|---|---|---|
| Age | 28.7 (9.4) | 24.7 (7.0) | 24.7 (5.1) | 28.4 (9.5) |
| Gender (% Female) | 593 (38.4) | 36 (30.3%) | 41 (34.5%) | 1,294 (37.3%) |
| Ethnicity | | | | |
| Asian | 115 (7.4%) | 10 (8.4%) | 16 (13.4%) | 265 (7.64%) |
| Black | 733 (47.4%) | 44 (37.0%) | 53 (44.5%) | 1,609 (46.4%) |
| Mixed | 67 (4.3%) | 7 (5.9%) | 4 (3.4%) | 125 (3.61%) |
| White | 454 (29.4%) | 35 (29.4%) | 31 (26.1%) | 980 (28.3%) |
| Other | 106 (6.9%) | 16 (13.4%) | 10 (8.4%) | 213 (6.14%) |
| Missing | 70 (4.5%) | 7 (5.9%) | 5 (4.2%) | 275 (7.93%) |
| Psychosis type | | | | |
| Affective | 218 (14.1%) | 7 (5.9%) | 11 (9.2%) | 398 (11.5%) |
| Non-affective | 1,261 (81.6%) | 100 (84.0%) | 102 (85.7%) | 2,875 (82.9%) |
| Postpartum psychosis | 1 (0.1%) | 0 (0%) | 0 (0%) | 7 (0.20%) |
| Substance-induced | 65 (4.2%) | 12 (10.1%) | 6 (5.0%) | 187 (5.39%) |
| Antidepressants | 450 (29.1%) | 44 (37.0%) | 39 (32.8%) | 749 (21.6%) |
| Antipsychotics | 1,182 (76.5%) | 81 (68.1%) | 77 (64.7%) | 2,935 (84.7%) |
| Mood stabilisers | 129 (8.3%) | 2 (1.7%) | 4 (3.4%) | 262 (7.56%) |

[1] Propensity score matched to detected group

**eTable 7:** Results Table for First Presentation

| Variable | Proportion DET+ | Proportion DET- | Odds Ratio | 95% Confidence Intervals | P value | P value corrected |
|---|---|---|---|---|---|---|
| Positive Symptoms | 0.68 | 0.66 | 1.1 | 0.74-1.66 | 0.634 | 0.82 |
| Negative Symptoms | 0.39 | 0.27 | 1.77 | 1.2-2.6 | 0.004 | 0.052 |
| Depressive Symptoms | 0.58 | 0.59 | 0.95 | 0.65-1.4 | 0.798 | 0.864 |
| Manic Symptoms | 0.51 | 0.53 | 0.92 | 0.63-1.33 | 0.648 | 0.82 |
| Disorganised Symptoms | 0.29 | 0.13 | 2.6 | 1.68-3.95 | <0.001 | <0.001 |
| Catatonic Symptoms | 0.01 | 0.04 | 0.22 | 0.01-0.99 | 0.13 | 0.416 |
| Other Symptoms | 0.92 | 0.87 | 1.59 | 0.86-3.3 | 0.169 | 0.418 |
| Substance Use | 0.45 | 0.44 | 1.04 | 0.71-1.51 | 0.835 | 0.886 |
| Agitation | 0.17 | 0.27 | 0.55 | 0.33-0.89 | 0.019 | 0.114 |
| Anergia | 0.03 | 0.01 | 4.93 | 1.33-14.98 | 0.008 | 0.083 |
| Anhedonia | 0.13 | 0.04 | 3.34 | 1.77-5.95 | <0.001 | <0.001 |
| Anxiety | 0.71 | 0.67 | 1.17 | 0.79-1.79 | 0.446 | 0.77 |
| Poor Appetite | 0.03 | 0.07 | 0.49 | 0.15-1.19 | 0.167 | 0.418 |
| Auditory Hallucinations | 0.19 | 0.24 | 0.74 | 0.46-1.17 | 0.22 | 0.52 |
| Bad Dreams | 0.02 | 0.01 | 1.42 | 0.22-5.02 | 0.644 | 0.82 |
| Blunted Affect | 0.09 | 0.06 | 1.65 | 0.81-3.06 | 0.137 | 0.416 |
| Cognitive Impairment | 0.75 | 0.68 | 1.38 | 0.91-2.14 | 0.144 | 0.416 |
| Concrete Thinking | 0 | 0.01 | NA | NA | NA | NA |
| Delusions | 0.24 | 0.28 | 0.78 | 0.5-1.2 | 0.277 | 0.6 |
| Disturbed Sleep | 0.35 | 0.38 | 0.87 | 0.59-1.28 | 0.49 | 0.772 |
| Diurnal Mood | 0 | 0 | NA | NA | NA | NA |
| Early Morning Wakening | 0 | 0.01 | NA | NA | NA | NA |
| Elation | 0.02 | 0.07 | 0.24 | 0.04-0.77 | 0.047 | 0.204 |

| | | | | | | |
|---|---|---|---|---|---|---|
| Emotionally Withdrawn | 0.19 | 0.13 | 1.58 | 0.96-2.51 | 0.063 | 0.252 |
| Grandiosity | 0.09 | 0.07 | 1.27 | 0.63-2.33 | 0.474 | 0.77 |
| Hallucination: Olfactory, tactile, or gustatory (OTG) | 0.01 | 0.02 | 0.42 | 0.02-2.01 | 0.4 | 0.743 |
| Feeling Helpless | 0.03 | 0.01 | 1.92 | 0.45-5.72 | 0.301 | 0.602 |
| Feeling Hopeless | 0.11 | 0.1 | 1.1 | 0.58-1.94 | 0.755 | 0.853 |
| Hostility | 0.03 | 0.1 | 0.25 | 0.06-0.66 | 0.018 | 0.114 |
| Poor Insight | 0.26 | 0.26 | 1.02 | 0.66-1.55 | 0.913 | 0.925 |
| Insomnia | 0.08 | 0.06 | 1.45 | 0.69-2.74 | 0.289 | 0.601 |
| Irritability | 0.16 | 0.19 | 0.82 | 0.48-1.33 | 0.436 | 0.77 |
| Lonely | 0.06 | 0.04 | 1.45 | 0.59-3.04 | 0.368 | 0.709 |
| Loss of Coherence | 0.02 | 0.05 | 0.36 | 0.06-1.16 | 0.156 | 0.418 |
| Mutism | 0.01 | 0.04 | 0.22 | 0.01-1.03 | 0.14 | 0.416 |
| Negative Symptoms | 0.07 | 0.03 | 2.5 | 1.06-5.2 | 0.022 | 0.114 |
| Nightmares | 0.05 | 0.04 | 1.19 | 0.45-2.6 | 0.694 | 0.82 |
| Paranoia | 0.51 | 0.48 | 1.15 | 0.79-1.67 | 0.472 | 0.77 |
| Passivity | 0.03 | 0.03 | 1.24 | 0.37-3.14 | 0.69 | 0.82 |
| Persecutory Delusions | 0.18 | 0.17 | 1.08 | 0.64-1.72 | 0.773 | 0.855 |
| Poor Motivation | 0.07 | 0.06 | 1.14 | 0.5-2.27 | 0.737 | 0.852 |
| Poverty of Speech | 0.03 | 0.02 | 1.09 | 0.26-3.1 | 0.886 | 0.921 |
| Poverty of Thought | 0.03 | 0.01 | 2.61 | 0.6-8.13 | 0.136 | 0.416 |
| Social Withdrawal | 0.1 | 0.05 | 1.94 | 0.98-3.54 | 0.042 | 0.199 |
| Stupor | 0 | 0 | NA | NA | NA | NA |
| Suicidal Ideation | 0.13 | 0.11 | 1.13 | 0.62-1.94 | 0.665 | 0.82 |
| Tangential Speech | 0.22 | 0.07 | 3.48 | 2.13-5.55 | <0.001 | <0.001 |
| Tearfulness | 0.17 | 0.19 | 0.88 | 0.52-1.41 | 0.606 | 0.82 |
| Thought Block | 0.08 | 0.04 | 2.29 | 1.07-4.43 | 0.021 | 0.114 |
| Thought Broadcast | 0.09 | 0.04 | 2.4 | 1.17-4.54 | 0.011 | 0.095 |
| Thought Insertion | 0.05 | 0.04 | 1.21 | 0.46-2.65 | 0.666 | 0.82 |
| Thought Withdrawal | 0.03 | 0.02 | 1.29 | 0.31-3.71 | 0.678 | 0.82 |

| | | | | | | |
|---|---|---|---|---|---|---|
| Visual Hallucination | 0.12 | 0.1 | 1.2 | 0.64-2.08 | 0.547 | 0.82 |
| Waxy Flexibility | 0 | 0 | NA | NA | NA | NA |
| Weight Loss | 0.13 | 0.09 | 1.38 | 0.75-2.37 | 0.269 | 0.6 |
| Feeling Worthless | 0.03 | 0.02 | 1.06 | 0.25-3 | 0.925 | 0.925 |
| Cannabis Use | 0.39 | 0.37 | 1.09 | 0.74-1.6 | 0.645 | 0.82 |
| Cocaine Use | 0.07 | 0.12 | 0.54 | 0.24 -1.05 | 0.096 | 0.378 |
| MDMA Use | 0.02 | 0.01 | 1.42 | 0.22 -5.02 | 0.644 | 0.819 |
| Tobacco Use | 0.19 | 0.19 | 1.05 | 0.64-1.67 | 0.826 | 0.880 |

| KEY | |
|---|---|
| | Clusters |
| | Features |
| | Significant Results |

**eTable 8:** Results Table Across Prodrome

| Variable | Mean DET+ (SD) | Mean DET- (SD) | Effect size (95% CIs) | P value | P value corrected |
|---|---|---|---|---|---|
| Positive Symptoms | 4.82 (5.86) | 4.30 (5.25) | 0.03 (0.0014-0.08) | 0.269 | 0.564 |
| Negative Symptoms | 2.37 (3.14) | 1.61 (2.53) | 0.09 (0.04-0.14) | 0.001 | 0.009 |
| Depressive Symptoms | 4.21 (4.76) | 3.66 (4.42) | 0.04 (0.0027-0.09) | 0.151 | 0.393 |
| Manic Symptoms | 3.46 (3.80) | 3.18 (4.05) | 0.03 (0.002-0.09) | 0.209 | 0.483 |
| Disorganised Symptoms | 1.02 (1.63) | 0.61 (1.26) | 0.09 (0.04-0.15) | 0.000 | 0.006 |
| Catatonic Symptoms | 0.19 (0.94) | 0.17 (0.68) | 0.02 (0.0016-0.07) | 0.395 | 0.661 |
| Other Symptoms | 7.14 (7.85) | 6.49 (7.13) | 0.04 (0.0025-0.09) | 0.129 | 0.369 |
| Substance Use | 2.57 (3.47) | 2.72 (4.68) | 0.03 (0.001-0.08) | 0.311 | 0.566 |
| Agitation | 1.23 (2.33) | 1.40 (2.32) | 0.04 (0.0025-0.09) | 0.177 | 0.425 |
| Anergia | 0.05 (0.23) | 0.03 (0.21) | 0.05 (0.0025-0.12) | 0.068 | 0.272 |
| Anhedonia | 0.55 (1.05) | 0.20 (0.63) | 0.12 (0.06-0.18) | <0.001 | <0.001 |
| Anxiety | 5.27 (5.82) | 4.69 (5.62) | 0.05 (0.0033-0.09) | 0.081 | 0.305 |
| Poor Appetite | 0.31 (0.77) | 0.28 (0.69) | 0.00 (0.00082-0.06) | 0.893 | 0.993 |
| Auditory Hallucinations | 1.30 (2.41) | 1.30 (2.16) | 0.02 (0.0012-0.07) | 0.396 | 0.661 |
| Bad Dreams | 0.07 (0.32) | 0.05 (0.27) | 0.02 (0.00093-0.08) | 0.492 | 0.738 |
| Blunted Affect | 0.40 (1.15) | 0.29 (0.75) | 0.01 (0.00084-0.07) | 0.655 | 0.873 |
| Cognitive Impairment | 5.34 (6.12) | 4.64 (5.37) | 0.04 (0.0028-0.09) | 0.114 | 0.351 |
| Concrete Thinking | 0.03 (0.21) | 0.04 (0.28) | 0.02 (0.0015-0.05) | 0.453 | 0.697 |
| Delusions | 1.47 (3.04) | 1.42 (2.21) | 0.03 (0.0011-0.08) | 0.290 | 0.564 |
| Disturbed Sleep | 2.44 (2.90) | 2.13 (2.95) | 0.04 (0.0025-0.09) | 0.117 | 0.351 |

| | | | | | |
|---|---|---|---|---|---|
| Diurnal Mood | 0.01 (0.09) | 0.01 (0.11) | 0.01 (0.0011-0.04) | 0.761 | 0.930 |
| Early Morning Wakening | 0.04 (0.19) | 0.04 (0.25) | 0.00 (0.001-0.06) | 0.981 | 0.993 |
| Elation | 0.20 (0.75) | 0.40 (1.25) | 0.05 (0.0055-0.09) | 0.064 | 0.272 |
| Emotionally Withdrawn | 0.87 (1.38) | 0.60 (1.19) | 0.08 (0.02-0.14) | 0.003 | 0.027 |
| Grandiosity | 0.24 (0.75) | 0.38 (1.17) | 0.03 (0.0012-0.07) | 0.336 | 0.593 |
| Hallucination: Olfactory, tactile, or gustatory (OTG) | 0.12 (0.58) | 0.10 (0.49) | 0.00 (0.00085-0.06) | 0.976 | 0.993 |
| Feeling Helpless | 0.10 (0.38) | 0.09 (0.38) | 0.00 (0.00078-0.06) | 0.983 | 0.993 |
| Feeling Hopeless | 0.74 (1.62) | 0.53 (1.26) | 0.04 (0.0023-0.09) | 0.138 | 0.376 |
| Hostility | 0.29 (1.09) | 0.56 (1.48) | 0.07 (0.03-0.11) | 0.005 | 0.039 |
| Poor Insight | 1.42 (1.98) | 1.49 (2.27) | 0.00 (0.00074-0.06) | 0.988 | 0.993 |
| Insomnia | 0.53 (1.17) | 0.31 (0.81) | 0.07 (0.01-0.13) | 0.010 | 0.065 |
| Irritability | 0.97 (1.73) | 1.08 (2.21) | 0.01 (0.00054-0.06) | 0.755 | 0.930 |
| Lonely | 0.27 (0.68) | 0.34 (1.06) | 0.01 (0.00061-0.06) | 0.800 | 0.941 |
| Loss of Coherence | 0.09 (0.37) | 0.19 (0.58) | 0.06 (0.02-0.1) | 0.029 | 0.177 |
| Mutism | 0.19 (0.94) | 0.16 (0.66) | 0.02 (0.0016-0.07) | 0.423 | 0.686 |
| Negative Symptoms | 0.49 (1.08) | 0.30 (0.98) | 0.09 (0.02-0.15) | 0.001 | 0.014 |
| Nightmares | 0.30 (0.73) | 0.25 (0.82) | 0.04 (0.0026-0.1) | 0.093 | 0.327 |
| Paranoia | 3.55 (4.49) | 2.84 (3.85) | 0.06 (0.0068-0.11) | 0.035 | 0.189 |
| Passivity | 0.19 (0.65) | 0.12 (0.52) | 0.05 (0.0043-0.12) | 0.055 | 0.272 |
| Persecutory Delusions | 0.99 (2.49) | 0.80 (1.48) | 0.02 (0.0012-0.08) | 0.450 | 0.697 |
| Poor Motivation | 0.71 (1.52) | 0.53 (1.30) | 0.03 (0.0013-0.09) | 0.266 | 0.564 |
| Poverty of Speech | 0.13 (0.73) | 0.09 (0.42) | 0.00 (0.00099-0.06) | 0.878 | 0.993 |
| Poverty of Thought | 0.12 (0.59) | 0.05 (0.38) | 0.03 (0.001-0.1) | 0.299 | 0.564 |

| | | | | | |
|---|---|---|---|---|---|
| Social Withdrawal | 0.41 (1.07) | 0.24 (0.74) | 0.04 (0.0022-0.1) | 0.169 | 0.421 |
| Stupor | 0.00 (0.00) | 0.01 (0.16) | 0.02 (0.0078-0.02) | 0.514 | 0.747 |
| Suicidal Ideation | 0.50 (0.90) | 0.61 (1.46) | 0.00 (0.00047-0.06) | 0.855 | 0.986 |
| Tangential Speech | 0.70 (1.42) | 0.35 (0.95) | 0.11 (0.04-0.17) | <0.001 | 0.001 |
| Tearfulness | 0.81 (1.56) | 0.91 (1.81) | 0.00 (0.00081-0.06) | 0.993 | 0.993 |
| Thought Block | 0.31 (0.89) | 0.17 (0.58) | 0.05 (0.0034-0.12) | 0.060 | 0.272 |
| Thought Broadcast | 0.29 (0.78) | 0.16 (0.62) | 0.07 (0.01-0.14) | 0.006 | 0.044 |
| Thought Insertion | 0.14 (0.40) | 0.14 (0.51) | 0.02 (0.0011-0.07) | 0.526 | 0.747 |
| Thought Withdrawal | 0.04 (0.21) | 0.06 (0.34) | 0.00 (0.00072-0.05) | 0.945 | 0.993 |
| Visual Hallucination | 0.38 (0.89) | 0.39 (0.99) | 0.01 (0.00094-0.06) | 0.775 | 0.930 |
| Waxy Flexibility | 0.00 (0.00) | 0.00 (0.09) | 0.01 (0.0072-0.02) | 0.681 | 0.888 |
| Weight Loss | 0.45 (1.01) | 0.46 (1.02) | 0.01 (0.0012-0.06) | 0.700 | 0.893 |
| Feeling Worthless | 0.19 (0.77) | 0.11 (0.45) | 0.02 (0.0011-0.08) | 0.539 | 0.747 |
| Cannabis Use | 2.11 (2.93) | 2.05 (3.82) | 0.04 (0.0035-0.09) | 0.103 | 0.345 |
| Cocaine Use | 0.50 (1.32) | 0.69 (2.35) | 0.02 (0.00087-0.07) | 0.548 | 0.747 |
| MDMA Use | 0.08 (0.38) | 0.06 (0.43) | 0.03 (0.0015-0.09) | 0.294 | 0.564 |
| Tobacco Use | 1.04 (1.72) | 1.12 (2.26) | 0.03 (0.0013-0.08) | 0.301 | 0.564 |

| KEY | |
|---|---|
| | Clusters |
| | Features |
| | Significant Results |

**eTable 9:** Results Table for First Presentation in Propensity Score Matched Sample

| Variable | Proportion DET+ | Proportion DET- | Odds Ratio | 95% Confidence Intervals | P value | P value corrected |
|---|---|---|---|---|---|---|
| Positive Symptoms | 0.69 | 0.68 | 0.96 | 0.56-1.66 | 0.889 | 1.00 |
| Negative Symptoms | 0.33 | 0.39 | 1.34 | 0.79-2.28 | 0.281 | 0.648 |
| Depressive Symptoms | 0.67 | 0.58 | 0.67 | 0.4-1.14 | 0.141 | 0.53 |
| Manic Symptoms | 0.57 | 0.51 | 0.79 | 0.47-1.31 | 0.363 | 0.74 |
| Disorganised Symptoms | 0.12 | 0.29 | 3.00 | 1.54-6.11 | 0.002 | 0.053 |
| Catatonic Symptoms | 0.05 | 0.01 | 0.16 | 0.01-0.95 | 0.092 | 0.488 |
| Other Symptoms | 0.91 | 0.92 | 1.11 | 0.45-2.77 | 0.819 | 1.00 |
| Substance Use | 0.54 | 0.45 | 0.71 | 0.43-1.19 | 0.195 | 0.53 |
| Agitation | 0.27 | 0.17 | 0.55 | 0.29-1.02 | 0.062 | 0.488 |
| Anergia | 0 | 0.03 | NA | NA | NA | NA |
| Anhedonia | 0.03 | 0.13 | 4.15 | 1.45-14.9 | 0.014 | 0.186 |
| Anxiety | 0.7 | 0.71 | 1.04 | 0.6-1.82 | 0.887 | 1.00 |
| Poor Appetite | 0.03 | 0.03 | 1.00 | 0.23-4.32 | 1.00 | 1.00 |
| Auditory Hallucinations | 0.29 | 0.19 | 0.57 | 0.31-1.04 | 0.072 | 0.488 |
| Bad Dreams | 0.03 | 0.02 | 0.66 | 0.09-4.06 | 0.653 | 1.00 |
| Blunted Affect | 0.06 | 0.09 | 1.63 | 0.62-4.57 | 0.331 | 0.702 |
| Cognitive Impairment | 0.76 | 0.75 | 0.96 | 0.53-1.72 | 0.881 | 1.00 |
| Concrete Thinking | 0 | 0 | NA | NA | NA | NA |
| Delusions | 0.32 | 0.24 | 0.66 | 0.37-1.16 | 0.149 | 0.53 |
| Disturbed Sleep | 0.45 | 0.35 | 0.68 | 0.4-1.14 | 0.146 | 0.53 |
| Diurnal Mood | 0.01 | 0 | NA | NA | NA | NA |
| Early Morning Wakening | 0 | 0 | NA | NA | NA | NA |
| Elation | 0.05 | 0.02 | 0.32 | 0.05-1.43 | 0.171 | 0.53 |

| | | | | | | |
|---|---|---|---|---|---|---|
| Emotionally Withdrawn | 0.2 | 0.19 | 0.95 | 0.5-1.8 | 0.871 | 1.00 |
| Grandiosity | 0.11 | 0.09 | 0.83 | 0.35-1.94 | 0.667 | 1.00 |
| Hallucination: Olfactory, tactile, or gustatory (OTG) | 0.03 | 0.01 | 0.24 | 0.01-1.68 | 0.21 | 0.53 |
| Feeling Helpless | 0.03 | 0.03 | 1.00 | 0.18-5.5 | 1.00 | 1.00 |
| Feeling Hopeless | 0.14 | 0.11 | 0.74 | 0.33-1.59 | 0.436 | 0.797 |
| Hostility | 0.13 | 0.03 | 0.17 | 0.04-0.52 | 0.005 | 0.088 |
| Poor Insight | 0.27 | 0.26 | 0.96 | 0.54-1.71 | 0.883 | 1.00 |
| Insomnia | 0.07 | 0.08 | 1.27 | 0.48-3.45 | 0.625 | 1.00 |
| Irritability | 0.16 | 0.16 | 1.00 | 0.5-2.01 | 1.00 | 1.00 |
| Lonely | 0.04 | 0.06 | 1.42 | 0.44-4.94 | 0.555 | 0.981 |
| Loss of Coherence | 0.03 | 0.02 | 0.49 | 0.07-2.57 | 0.417 | 0.789 |
| Mutism | 0.05 | 0.01 | 0.16 | 0.01-0.95 | 0.092 | 0.488 |
| Negative Symptoms | 0.04 | 0.07 | 1.64 | 0.53-5.58 | 0.396 | 0.777 |
| Nightmares | 0.06 | 0.05 | 0.85 | 0.27-2.63 | 0.776 | 1.00 |
| Paranoia | 0.53 | 0.51 | 0.93 | 0.56-1.56 | 0.795 | 1.00 |
| Passivity | 0.03 | 0.03 | 1.34 | 0.29-6.95 | 0.702 | 1.00 |
| Persecutory Delusions | 0.16 | 0.18 | 1.13 | 0.57-2.24 | 0.729 | 1.00 |
| Poor Motivation | 0.07 | 0.07 | 1.00 | 0.36-2.81 | 1.00 | 1.00 |
| Poverty of Speech | 0.06 | 0.03 | 0.41 | 0.09-1.53 | 0.209 | 0.53 |
| Poverty of Thought | 0.02 | 0.03 | 1.51 | 0.25-11.64 | 0.653 | 1.00 |
| Social Withdrawal | 0.04 | 0.1 | 2.56 | 0.92-8.26 | 0.087 | 0.488 |
| Stupor | 0 | 0 | NA | NA | NA | NA |
| Suicidal Ideation | 0.08 | 0.13 | 1.76 | 0.75-4.36 | 0.201 | 0.53 |
| Tangential Speech | 0.07 | 0.22 | 3.88 | 1.75-9.54 | 0.002 | 0.053 |
| Tearfulness | 0.24 | 0.17 | 0.66 | 0.34-1.24 | 0.198 | 0.53 |
| Thought Block | 0.04 | 0.08 | 2.09 | 0.72-6.9 | 0.191 | 0.53 |
| Thought Broadcast | 0.03 | 0.09 | 2.93 | 0.97-10.82 | 0.073 | 0.488 |
| Thought Insertion | 0.04 | 0.05 | 1.21 | 0.35-4.31 | 0.758 | 1.00 |
| Thought Withdrawal | 0.03 | 0.03 | 0.74 | 0.14-3.44 | 0.702 | 1.00 |

| | | | | | | |
|---|---|---|---|---|---|---|
| Visual Hallucination | 0.12 | 0.12 | 1.00 | 0.45-2.22 | 1.00 | 1.00 |
| Waxy Flexibility | 0.01 | 0 | NA | NA | NA | NA |
| Weight Loss | 0.11 | 0.13 | 1.18 | 0.53-2.63 | 0.688 | 1.00 |
| Feeling Worthless | 0.05 | 0.03 | 0.49 | 0.1-1.89 | 0.317 | 0.7 |
| Cannabis Use | 0.46 | 0.39 | 0.73 | 0.44-1.23 | 0.238 | 0.573 |
| Cocaine Use | 0.13 | 0.07 | 0.5 | 0.19-1.2 | 0.13 | 0.53 |
| MDMA Use | 0 | 0.02 | NA | NA | NA | NA |
| Tobacco Use | 0.19 | 0.19 | 1.00 | 0.52-1.91 | 1.00 | 1.00 |

| KEY | |
|---|---|
| | Clusters |
| | Features |
| | Significant Results |

**eTable 10:** Results Table Across Prodrome in Propensity Score Matched Sample

| Variable | Mean DET+ (SD) | Mean DET-(SD) | Effect size (95% CIs) | P value | P value corrected |
|---|---|---|---|---|---|
| Positive Symptoms | 4.82 (5.86) | 3.50 (3.78) | 0.11 (0.0086-0.24) | 0.110 | 0.309 |
| Negative Symptoms | 2.37 (3.14) | 1.46 (2.24) | 0.20 (0.08-0.33) | 0.003 | 0.042 |
| Depressive Symptoms | 4.21 (4.76) | 3.04 (3.70) | 0.15 (0.03-0.27) | 0.021 | 0.127 |
| Manic Symptoms | 3.46 (3.80) | 2.39 (3.00) | 0.16 (0.03-0.29) | 0.014 | 0.102 |
| Disorganised Symptoms | 1.02 (1.63) | 0.53 (1.10) | 0.19 (0.06-0.32) | 0.004 | 0.042 |
| Catatonic Symptoms | 0.19 (0.94) | 0.19 (0.76) | 0.06 (0.004-0.18) | 0.364 | 0.591 |
| Other Symptoms | 7.14 (7.85) | 5.66 (6.12) | 0.13 (0.01-0.26) | 0.053 | 0.197 |
| Substance Use | 2.57 (3.47) | 2.50 (3.99) | 0.07 (0.0039-0.19) | 0.289 | 0.549 |
| Agitation | 1.23 (2.33) | 1.16 (1.71) | 0.05 (0.0036-0.18) | 0.477 | 0.722 |
| Anergia | 0.05 (0.23) | 0.04 (0.30) | 0.09 (0.0053-0.2) | 0.157 | 0.392 |
| Anhedonia | 0.55 (1.05) | 0.16 (0.49) | 0.22 (0.1-0.34) | 0.001 | 0.042 |
| Anxiety | 5.27 (5.82) | 4.07 (4.91) | 0.14 (0.02-0.27) | 0.038 | 0.167 |
| Poor Appetite | 0.31 (0.77) | 0.27 (0.69) | 0.03 (0.0015-0.15) | 0.697 | 0.823 |
| Auditory Hallucinations | 1.30 (2.41) | 1.12 (1.65) | 0.03 (0.0021-0.17) | 0.601 | 0.787 |
| Bad Dreams | 0.07 (0.32) | 0.06 (0.31) | 0.02 (0.0022-0.15) | 0.764 | 0.860 |
| Blunted Affect | 0.40 (1.15) | 0.20 (0.48) | 0.05 (0.003-0.18) | 0.441 | 0.685 |
| Cognitive Impairment | 5.34 (6.12) | 4.38 (5.08) | 0.11 (0.0067-0.24) | 0.108 | 0.309 |
| Concrete Thinking | 0.03 (0.21) | 0.05 (0.26) | 0.08 (0.0038-0.19) | 0.256 | 0.521 |
| Delusions | 1.47 (3.04) | 1.14 (1.68) | 0.00 (0.0018-0.14) | 0.948 | 0.996 |
| Disturbed Sleep | 2.44 (2.90) | 1.75 (2.32) | 0.14 | 0.040 | 0.167 |

| | | | | | |
|---|---|---|---|---|---|
| | | | (0.01-0.28) | | |
| Diurnal Mood | 0.01 (0.09) | 0.02 (0.13) | 0.04 (0.0011-0.14) | 0.565 | 0.780 |
| Early Morning Wakening | 0.04 (0.19) | 0.05 (0.35) | 0.02 (0.0019-0.15) | 0.724 | 0.838 |
| Elation | 0.20 (0.75) | 0.31 (0.84) | 0.11 (0.01-0.24) | 0.103 | 0.309 |
| Emotionally Withdrawn | 0.87 (1.38) | 0.65 (1.25) | 0.11 (0.0063-0.24) | 0.102 | 0.309 |
| Grandiosity | 0.24 (0.75) | 0.35 (0.84) | 0.09 (0.0078-0.22) | 0.160 | 0.392 |
| Hallucination: Olfactory, tactile, or gustatory (OTG) | 0.12 (0.58) | 0.03 (0.16) | 0.09 (0.0068-0.2) | 0.192 | 0.435 |
| Feeling Helpless | 0.10 (0.38) | 0.11 (0.36) | 0.03 (0.0023-0.16) | 0.647 | 0.812 |
| Feeling Hopeless | 0.74 (1.62) | 0.37 (0.88) | 0.12 (0.02-0.25) | 0.061 | 0.210 |
| Hostility | 0.29 (1.09) | 0.42 (0.88) | 0.14 (0.03-0.26) | 0.030 | 0.159 |
| Poor Insight | 1.42 (1.98) | 1.23 (1.74) | 0.04 (0.0029-0.17) | 0.574 | 0.780 |
| Insomnia | 0.53 (1.17) | 0.18 (0.66) | 0.21 (0.1-0.33) | 0.002 | 0.042 |
| Irritability | 0.97 (1.73) | 0.81 (1.23) | 0.00 (0.0032-0.14) | 0.978 | 0.996 |
| Lonely | 0.27 (0.68) | 0.35 (0.83) | 0.06 (0.0031-0.19) | 0.371 | 0.591 |
| Loss of Coherence | 0.09 (0.37) | 0.19 (0.51) | 0.13 (0.01-0.25) | 0.053 | 0.197 |
| Mutism | 0.19 (0.94) | 0.19 (0.76) | 0.06 (0.0032-0.19) | 0.364 | 0.591 |
| Negative Symptoms | 0.49 (1.08) | 0.19 (0.72) | 0.20 (0.07-0.32) | 0.002 | 0.042 |
| Nightmares | 0.30 (0.73) | 0.22 (0.64) | 0.07 (0.0035-0.2) | 0.288 | 0.549 |
| Paranoia | 3.55 (4.49) | 2.14 (2.61) | 0.17 (0.04-0.3) | 0.012 | 0.102 |
| Passivity | 0.19 (0.65) | 0.04 (0.25) | 0.16 (0.04-0.27) | 0.014 | 0.102 |
| Persecutory Delusions | 0.99 (2.49) | 0.55 (0.95) | 0.02 (0.0018-0.15) | 0.806 | 0.865 |
| Poor Motivation | 0.71 (1.52) | 0.43 (1.01) | 0.09 (0.0075-0.21) | 0.167 | 0.395 |
| Poverty of Speech | 0.13 (0.73) | 0.14 (0.73) | 0.02 (0.0024-0.16) | 0.797 | 0.865 |
| Poverty of Thought | 0.12 (0.59) | 0.02 (0.13) | 0.10 | 0.146 | 0.392 |

| | | | | | |
|---|---|---|---|---|---|
| | | | (0.0066-0.2) | | |
| Social Withdrawal | 0.41 (1.07) | 0.28 (0.94) | 0.08 (0.005-0.21) | 0.208 | 0.455 |
| Stupor | 0.00 (0.00) | 0.00 (0.00) | NA (NA) | NA | NA |
| Suicidal Ideation | 0.50 (0.90) | 0.50 (1.41) | 0.04 (0.0027-0.18) | 0.516 | 0.760 |
| Tangential Speech | 0.70 (1.42) | 0.37 (0.94) | 0.16 (0.03-0.28) | 0.016 | 0.102 |
| Tearfulness | 0.81 (1.56) | 0.87 (1.82) | 0.03 (0.0015-0.17) | 0.628 | 0.805 |
| Thought Block | 0.31 (0.89) | 0.09 (0.32) | 0.14 (0.02-0.26) | 0.032 | 0.160 |
| Thought Broadcast | 0.29 (0.78) | 0.18 (0.45) | 0.06 (0.0033-0.19) | 0.348 | 0.591 |
| Thought Insertion | 0.14 (0.40) | 0.12 (0.37) | 0.04 (0.0032-0.17) | 0.535 | 0.769 |
| Thought Withdrawal | 0.04 (0.21) | 0.04 (0.21) | 0.00 (0.0017-0.15) | 1.000 | 1.000 |
| Visual Hallucination | 0.38 (0.89) | 0.34 (0.69) | 0.03 (0.0015-0.17) | 0.661 | 0.812 |
| Waxy Flexibility | 0.00 (0.00) | 0.03 (0.28) | 0.07 (0.06-0.13) | 0.322 | 0.591 |
| Weight Loss | 0.45 (1.01) | 0.40 (0.92) | 0.06 (0.0038-0.18) | 0.359 | 0.591 |
| Feeling Worthless | 0.19 (0.77) | 0.10 (0.40) | 0.04 (0.0022-0.16) | 0.582 | 0.780 |
| Cannabis Use | 2.11 (2.93) | 1.96 (3.41) | 0.08 (0.0044-0.2) | 0.243 | 0.513 |
| Cocaine Use | 0.50 (1.32) | 0.52 (1.89) | 0.02 (0.0021-0.15) | 0.773 | 0.860 |
| MDMA Use | 0.08 (0.38) | 0.12 (0.58) | 0.00 (0.0017-0.14) | 0.979 | 0.996 |
| Tobacco Use | 1.04 (1.72) | 1.09 (1.81) | 0.03 (0.0033-0.15) | 0.680 | 0.819 |

| KEY | |
|---|---|
| | Clusters |
| | Features |
| | Significant Results |

**eTable 11:** Results Table Across Prodrome in Sample with 1-month Antecedent Period

| Variable | Proportion DET+ | Proportion DET- | Odds Ratio | 95% Confidence Intervals | P value | P value corrected |
|---|---|---|---|---|---|---|
| Positive Symptoms | 0.73 | 0.76 | 0.87 | 0.62-1.23 | 0.417 | 0.571 |
| Negative Symptoms | 0.44 | 0.32 | 1.62 | 1.19-2.2 | 0.002 | 0.017 |
| Depressive Symptoms | 0.65 | 0.65 | 0.99 | 0.72-1.37 | 0.944 | 0.944 |
| Manic Symptoms | 0.58 | 0.62 | 0.85 | 0.62-1.16 | 0.294 | 0.525 |
| Disorganised Symptoms | 0.3 | 0.22 | 1.54 | 1.1-2.14 | 0.011 | 0.052 |
| Catatonic Symptoms | 0.03 | 0.07 | 0.36 | 0.13-0.8 | 0.026 | 0.097 |
| Other Symptoms | 0.93 | 0.89 | 1.5 | 0.87-2.8 | 0.173 | 0.375 |
| Substance Use | 0.48 | 0.51 | 0.91 | 0.67-1.24 | 0.55 | 0.681 |
| Agitation | 0.2 | 0.36 | 0.45 | 0.3-0.64 | <0.001 | <0.001 |
| Anergia | 0.03 | 0.01 | 3.94 | 1.29-9.93 | 0.007 | 0.036 |
| Anhedonia | 0.12 | 0.05 | 2.73 | 1.65-4.34 | <0.001 | <0.001 |
| Anxiety | 0.74 | 0.71 | 1.17 | 0.84-1.67 | 0.366 | 0.566 |
| Poor Appetite | 0.04 | 0.08 | 0.52 | 0.23-1 | 0.075 | 0.195 |
| Auditory Hallucinations | 0.24 | 0.33 | 0.63 | 0.44-0.89 | 0.012 | 0.052 |
| Bad Dreams | 0.01 | 0.01 | 0.81 | 0.13-2.69 | 0.776 | 0.824 |
| Blunted Affect | 0.09 | 0.09 | 1.02 | 0.58-1.69 | 0.937 | 0.944 |
| Cognitive Impairment | 0.78 | 0.74 | 1.25 | 0.88-1.83 | 0.229 | 0.433 |
| Concrete Thinking | 0 | 0.01 | NA | NA | NA | NA |
| Delusions | 0.28 | 0.39 | 0.6 | 0.42-0.83 | 0.003 | 0.022 |
| Disturbed Sleep | 0.43 | 0.47 | 0.87 | 0.64-1.18 | 0.381 | 0.566 |
| Diurnal Mood | 0 | 0 | NA | NA | NA | NA |
| Early Morning Wakening | 0 | 0.01 | NA | NA | NA | NA |
| Elation | 0.04 | 0.11 | 0.34 | 0.14-0.67 | 0.005 | 0.029 |

| | | | | | | |
|---|---|---|---|---|---|---|
| Emotionally Withdrawn | 0.26 | 0.16 | 1.83 | 1.28-2.58 | 0.001 | 0.013 |
| Grandiosity | 0.1 | 0.13 | 0.73 | 0.42-1.19 | 0.233 | 0.433 |
| Hallucination: Olfactory, tactile, or gustatory (OTG) | 0.02 | 0.04 | 0.44 | 0.11-1.2 | 0.17 | 0.375 |
| Feeling Helpless | 0.03 | 0.02 | 2.13 | 0.8-4.72 | 0.088 | 0.208 |
| Feeling Hopeless | 0.09 | 0.11 | 0.81 | 0.46-1.33 | 0.43 | 0.573 |
| Hostility | 0.06 | 0.14 | 0.41 | 0.21-0.72 | 0.005 | 0.029 |
| Poor Insight | 0.28 | 0.31 | 0.85 | 0.6-1.18 | 0.347 | 0.564 |
| Insomnia | 0.09 | 0.07 | 1.32 | 0.75-2.2 | 0.303 | 0.525 |
| Irritability | 0.2 | 0.26 | 0.7 | 0.47-1.01 | 0.063 | 0.172 |
| Lonely | 0.05 | 0.05 | 1.12 | 0.52-2.13 | 0.741 | 0.803 |
| Loss of Coherence | 0.04 | 0.09 | 0.5 | 0.22-0.97 | 0.061 | 0.172 |
| Mutism | 0.03 | 0.07 | 0.37 | 0.13-0.81 | 0.029 | 0.101 |
| Negative Symptoms | 0.07 | 0.04 | 2.08 | 1.09-3.67 | 0.016 | 0.064 |
| Nightmares | 0.07 | 0.05 | 1.47 | 0.76-2.62 | 0.215 | 0.43 |
| Paranoia | 0.58 | 0.58 | 0.98 | 0.72-1.34 | 0.918 | 0.944 |
| Passivity | 0.03 | 0.04 | 0.8 | 0.31-1.7 | 0.601 | 0.72 |
| Persecutory Delusions | 0.2 | 0.24 | 0.78 | 0.53-1.13 | 0.209 | 0.43 |
| Poor Motivation | 0.07 | 0.07 | 1.12 | 0.6-1.94 | 0.697 | 0.771 |
| Poverty of Speech | 0.02 | 0.04 | 0.6 | 0.18-1.44 | 0.315 | 0.528 |
| Poverty of Thought | 0.03 | 0.01 | 2.33 | 0.79-5.54 | 0.083 | 0.206 |
| Social Withdrawal | 0.12 | 0.06 | 2.1 | 1.28-3.31 | 0.002 | 0.017 |
| Stupor | 0 | 0 | NA | NA | NA | NA |
| Suicidal Ideation | 0.12 | 0.15 | 0.82 | 0.51-1.27 | 0.403 | 0.566 |
| Tangential Speech | 0.21 | 0.13 | 1.86 | 1.26-2.68 | 0.001 | 0.013 |
| Tearfulness | 0.2 | 0.22 | 0.85 | 0.57-1.22 | 0.392 | 0.566 |
| Thought Block | 0.09 | 0.08 | 1.18 | 0.67-1.96 | 0.536 | 0.68 |
| Thought Broadcast | 0.1 | 0.06 | 1.68 | 0.96-2.77 | 0.052 | 0.159 |
| Thought Insertion | 0.04 | 0.06 | 0.73 | 0.32-1.41 | 0.386 | 0.566 |
| Thought Withdrawal | 0.03 | 0.02 | 1.37 | 0.52-2.95 | 0.473 | 0.615 |

| | | | | | | |
|---|---|---|---|---|---|---|
| Visual Hallucination | 0.15 | 0.14 | 1.12 | 0.72-1.68 | 0.609 | 0.72 |
| Waxy Flexibility | 0 | 0 | NA | NA | NA | NA |
| Weight Loss | 0.17 | 0.12 | 1.53 | 1-2.26 | 0.039 | 0.127 |
| Feeling Worthless | 0.03 | 0.03 | 1.23 | 0.47-2.64 | 0.633 | 0.731 |
| Cannabis Use | 0.4 | 0.42 | 0.94 | 0.69-1.28 | 0.691 | 0.771 |
| Cocaine Use | 0.08 | 0.13 | 0.59 | 0.33-0.99 | 0.28 | 0.173 |
| MDMA Use | 0.02 | 0.02 | 0.91 | 0.22-2.53 | 0.882 | 0.933 |
| Tobacco Use | 0.22 | 0.24 | 0.88 | 0.6-1.25 | 0.485 | 0.635 |

| KEY | |
|---|---|
| | Clusters |
| | Features |
| | Significant Results |

**eTable 12:** Results Table Across Prodrome in Sample with 1-month Antecedent Period

| Variable | Mean DET+ (SD) | Mean DET- (SD) | Effect size (95% CIs) | P value | P value corrected |
|---|---|---|---|---|---|
| Positive Symptoms | 5.08 (5.65) | 4.09 (4.91) | 0.06 (0.01-0.09) | 0.004 | 0.012 |
| Negative Symptoms | 2.51 (3.16) | 1.56 (2.36) | 0.10 (0.06-0.14) | <0.001 | <0.001 |
| Depressive Symptoms | 4.47 (4.89) | 3.35 (4.05) | 0.07 (0.03-0.11) | <0.001 | 0.001 |
| Manic Symptoms | 3.76 (4.04) | 3.02 (3.75) | 0.05 (0.01-0.09) | 0.005 | 0.017 |
| Disorganised Symptoms | 1.14 (1.66) | 0.76 (1.32) | 0.07 (0.03-0.11) | <0.001 | 0.002 |
| Catatonic Symptoms | 0.25 (1.09) | 0.24 (0.81) | 0.02 (0.00081-0.05) | 0.398 | 0.519 |
| Other Symptoms | 7.24 (7.85) | 5.61 (6.91) | 0.08 (0.04-0.11) | <0.001 | <0.001 |
| Substance Use | 2.81 (3.95) | 2.55 (4.43) | 0.03 (0.0026-0.07) | 0.068 | 0.131 |
| Agitation | 1.39 (2.26) | 1.49 (2.21) | 0.03 (0.0011-0.06) | 0.176 | 0.278 |
| Anergia | 0.04 (0.21) | 0.03 (0.21) | 0.04 (0.0022-0.09) | 0.056 | 0.116 |
| Anhedonia | 0.54 (1.03) | 0.19 (0.59) | 0.12 (0.07-0.17) | <0.001 | <0.001 |
| Anxiety | 5.55 (5.83) | 4.14 (5.36) | 0.08 (0.04-0.12) | <0.001 | <0.001 |
| Poor Appetite | 0.34 (0.73) | 0.27 (0.65) | 0.02 (0.00095-0.06) | 0.237 | 0.347 |
| Auditory Hallucinations | 1.43 (2.22) | 1.38 (2.11) | 0.01 (0.00062-0.05) | 0.666 | 0.783 |
| Bad Dreams | 0.05 (0.27) | 0.06 (0.30) | 0.01 (0.00075-0.04) | 0.755 | 0.783 |
| Blunted Affect | 0.45 (1.08) | 0.36 (0.83) | 0.01 (0.00075-0.04) | 0.727 | 0.783 |
| Cognitive Impairment | 5.53 (6.11) | 4.17 (5.10) | 0.07 (0.04-0.11) | <0.001 | 0.001 |
| Concrete Thinking | 0.03 (0.22) | 0.04 (0.28) | 0.02 (0.0012-0.04) | 0.323 | 0.447 |
| Delusions | 1.55 (2.68) | 1.58 (2.14) | 0.03 (0.0018-0.07) | 0.116 | 0.204 |
| Disturbed Sleep | 2.76 (2.99) | 2.06 (2.71) | 0.07 | <0.001 | 0.002 |

| | | | | | |
|---|---|---|---|---|---|
| | | | (0.03-0.11) | | |
| Diurnal Mood | 0.01 (0.11) | 0.01 (0.12) | 0.00 (0.00067-0.04) | 0.974 | 0.974 |
| Early Morning Wakening | 0.02 (0.15) | 0.03 (0.21) | 0.01 (0.00066-0.04) | 0.741 | 0.783 |
| Elation | 0.24 (0.78) | 0.45 (1.24) | 0.05 (0.02-0.08) | 0.008 | 0.022 |
| Emotionally Withdrawn | 1.06 (1.52) | 0.63 (1.21) | 0.09 (0.05-0.13) | <0.001 | <0.001 |
| Grandiosity | 0.29 (0.78) | 0.46 (1.19) | 0.04 (0.0079-0.07) | 0.045 | 0.104 |
| Hallucination: Olfactory, tactile, or gustatory (OTG) | 0.12 (0.54) | 0.12 (0.51) | 0.01 (0.00063-0.04) | 0.733 | 0.783 |
| Feeling Helpless | 0.13 (0.48) | 0.08 (0.34) | 0.02 (0.0013-0.07) | 0.218 | 0.327 |
| Feeling Hopeless | 0.76 (1.54) | 0.49 (1.17) | 0.06 (0.02-0.1) | 0.003 | 0.012 |
| Hostility | 0.38 (1.18) | 0.64 (1.55) | 0.06 (0.03-0.1) | 0.001 | 0.003 |
| Poor Insight | 1.69 (1.99) | 1.44 (2.13) | 0.04 (0.0083-0.08) | 0.020 | 0.049 |
| Insomnia | 0.54 (1.17) | 0.30 (0.79) | 0.07 (0.02-0.11) | <0.001 | 0.002 |
| Irritability | 1.11 (1.82) | 1.17 (2.14) | 0.01 (0.00062-0.05) | 0.707 | 0.783 |
| Lonely | 0.29 (0.81) | 0.29 (0.94) | 0.01 (0.00095-0.05) | 0.537 | 0.644 |
| Loss of Coherence | 0.17 (0.48) | 0.27 (0.68) | 0.04 (0.0039-0.07) | 0.050 | 0.109 |
| Mutism | 0.25 (1.09) | 0.24 (0.80) | 0.02 (0.00083-0.05) | 0.422 | 0.539 |
| Negative Symptoms | 0.49 (1.20) | 0.28 (0.95) | 0.10 (0.05-0.14) | <0.001 | <0.001 |
| Nightmares | 0.35 (0.84) | 0.22 (0.79) | 0.06 (0.02-0.11) | 0.001 | 0.003 |
| Paranoia | 3.82 (4.44) | 2.84 (3.61) | 0.06 (0.02-0.1) | 0.001 | 0.003 |
| Passivity | 0.21 (0.76) | 0.14 (0.63) | 0.03 (0.0015-0.08) | 0.094 | 0.177 |
| Persecutory Delusions | 1.02 (2.20) | 0.91 (1.46) | 0.02 (0.00099-0.06) | 0.203 | 0.313 |
| Poor Motivation | 0.66 (1.43) | 0.44 (1.13) | 0.04 (0.006-0.08) | 0.018 | 0.048 |
| Poverty of Speech | 0.16 (0.84) | 0.12 (0.46) | 0.00 (0.00058-0.04) | 0.969 | 0.974 |
| Poverty of Thought | 0.12 (0.52) | 0.06 (0.36) | 0.04 | 0.061 | 0.122 |

| | | | | | |
|---|---|---|---|---|---|
| | | | (0.0015-0.09) | | |
| Social Withdrawal | 0.47 (1.01) | 0.25 (0.72) | 0.08 (0.04-0.12) | <0.001 | <0.001 |
| Stupor | 0.00 (0.00) | 0.01 (0.13) | 0.02 (0.01-0.02) | 0.327 | 0.447 |
| Suicidal Ideation | 0.61 (1.08) | 0.58 (1.36) | 0.02 (0.001-0.06) | 0.342 | 0.456 |
| Tangential Speech | 0.78 (1.43) | 0.45 (1.00) | 0.07 (0.03-0.12) | <0.001 | 0.001 |
| Tearfulness | 1.03 (1.73) | 0.88 (1.65) | 0.03 (0.0013-0.07) | 0.171 | 0.278 |
| Thought Block | 0.35 (0.93) | 0.23 (0.67) | 0.03 (0.0021-0.07) | 0.128 | 0.219 |
| Thought Broadcast | 0.31 (0.78) | 0.19 (0.66) | 0.05 (0.0093-0.1) | 0.006 | 0.019 |
| Thought Insertion | 0.18 (0.71) | 0.18 (0.64) | 0.01 (7e-04-0.04) | 0.757 | 0.783 |
| Thought Withdrawal | 0.11 (0.42) | 0.07 (0.46) | 0.03 (0.0015-0.07) | 0.112 | 0.203 |
| Visual Hallucination | 0.49 (0.95) | 0.45 (1.01) | 0.01 (0.00092-0.05) | 0.706 | 0.783 |
| Waxy Flexibility | 0.00 (0.00) | 0.00 (0.09) | 0.01 (0.0084-0.02) | 0.460 | 0.563 |
| Weight Loss | 0.62 (1.15) | 0.48 (1.00) | 0.04 (0.0029-0.08) | 0.042 | 0.101 |
| Feeling Worthless | 0.21 (0.69) | 0.10 (0.43) | 0.05 (0.0077-0.1) | 0.011 | 0.031 |
| Cannabis Use | 2.34 (3.58) | 1.97 (3.73) | 0.04 (0.0029-0.08) | 0.051 | 0.109 |
| Cocaine Use | 0.46 (1.23) | 0.64 (2.16) | 0.03 (0.0017-0.06) | 0.157 | 0.261 |
| MDMA Use | 0.10 (0.48) | 0.07 (0.47) | 0.01 (0.00085-0.06) | 0.457 | 0.563 |
| Tobacco Use | 1.22 (1.90) | 1.15 (2.21) | 0.02 (0.00087-0.06) | 0.270 | 0.386 |

| KEY | |
|---|---|
| | Clusters |
| | Features |
| | Significant Results |
